# Oxidative stress and genetic susceptibility to cross-reactive and selective NSAID hypersensitivity

**DOI:** 10.64898/2026.01.29.26345114

**Authors:** JM García-Menaya, J Gómez-Tabales, S Ladera-Navarro, M Martí, N Blanca-López, JAG Agúndez, E García-Martín, P Ayuso

**Affiliations:** University Institute of Molecular Pathology Biomarkers, Universidad de Extremadura, Cáceres, Spain; Allergy Service, Badajoz University Hospital, Badajoz, Spain; Allergy Service, Infanta Leonor University Hospital, Madrid, Spain

**Keywords:** redox, drug hypersensitivity reactions, non-steroidal anti-inflammatory drugs, cross-reactive intolerance, single-NSAID–induced urticaria/angioedema/anaphylaxis, single nucleotide variants

## Abstract

**Aims:** Nonsteroidal anti inflammatory drugs (NSAIDs) are a leading cause of drug hypersensitivity reactions (HRs), yet genetic determinants of individual susceptibility remain unclear. Growing evidence implicates oxidative stress in these reactions. This study aimed to identify genetic variants in redox and immune related pathways associated with cross reactive NSAID hypersensitivity (CR NSAIDs) and single NSAID induced urticaria/angioedema/anaphylaxis (SNIUAA), and to characterize their functional relevance.

**Results:** Genetic association analyses identified significant associations between NSAID HRs and single nucleotide variants (SNVs) in genes involved in redox homeostasis. In CR NSAIDs patients, SNV *GSTM5* rs11101989 and *GSTZ1* haplotype (rs1046428, rs7975, rs7972) were associated with increased risk of developing HRs, suggesting impaired glutathione dependent detoxification as the underlying factor. Functional analyses demonstrated that carriers of the *GSTM5* rs11101989 CC genotype had significantly reduced serum GST activity. In SNIUAA patients, *SOD1* variants (rs2070424 and rs2833483) were consistently associated with increased susceptibility and reduced serum superoxide dismutase activity. Additionally, *AKR1C3* rs34186955 is also associated with increased susceptibility whereas the variants *AKR1C3* rs12529 and *IL4R* rs1805016 are associated with decreased susceptibility, highlighting the interplay between prostaglandin metabolism, redox regulation, and immune signalling.

**Innovation:** Integration of genetic and functional enzymatic validation provides mechanistic evidence that oxidative stress pathways modulate distinct NSAID hypersensitivity phenotypes. Identification of *GSTM5* and *SOD1* variants with measurable functional impact establishes promising biomarkers to guide personalized risk assessment.

**Conclusion:** Genetic variability in oxidative⍰stress and immune regulatory pathways influences susceptibility to cross reactive and selective NSAID hypersensitivity, supporting biomarker based strategies for safer NSAID prescribing.

## 1. Introduction

Nonsteroidal anti-inflammatory drugs (NSAIDs) constitute one of the most widely utilized pharmacological classes for their anti-inflammatory, analgesic, and antipyretic properties (Gómez-Acebo et al., 2018; Hessami et al., 2025). However, their extensive use is associated with hypersensitivity reactions (HRs), which represent a significant proportion of adverse drug reactions (ADRs) and impose a considerable burden on patients and healthcare systems (Blanca-Lopez et al., 2019). Epidemiological data indicate that the prevalence of NSAID-induced HRs ranges from 1.5% to 3% in the general population and may reach up to 30% among individuals with comorbid conditions such as asthma, chronic rhinosinusitis with nasal polyps, or chronic spontaneous urticaria (Sousa-Pinto et al., 2017) (Sánchez et al., 2019; Sánchez-Borges et al., 2015).

NSAID hypersensitivity encompasses a broad spectrum of clinical presentations, from isolated cutaneous symptoms to severe, multisystem involvement, and may arise through distinct immunological and non-immunological mechanisms (Kowalski et al., 2011). The European Academy of Allergy and Clinical Immunology (EAACI) classifies these reactions into two major categories: (i) cross-reactive hypersensitivity reactions (CRs), typically associated with cyclooxygenase (COX) inhibition and dysregulated arachidonic acid metabolism, and (ii) selective reactions (SRs), which are mediated by specific immune mechanisms and occur in response to a single NSAID or a chemically related group, while other structurally unrelated NSAIDs are tolerated (Kowalski et al., 2011). More recently, the World Allergy Organization (WAO) proposed an updated classification system that delineates immediate HRs into four principal phenotypes: NSAID-exacerbated respiratory disease (NERD), NSAID-exacerbated cutaneous disease (NECD), NSAID-induced urticaria/angioedema/anaphylaxis (NIUAA), and single-NSAID–induced urticaria/angioedema/anaphylaxis (SNIUAA). Delayed reactions, although less frequent, are defined as single-NSAID–induced delayed hypersensitivity reactions (SNIDHRs), which involve T-cell–mediated mechanisms (Romano et al., 2025). This framework also acknowledges mixed phenotypes, such as NERD patients presenting with cutaneous symptoms or NECD patients exhibiting respiratory features, and incorporates NSAID-exacerbated food allergy, highlighting the role of NSAIDs as cofactors in allergic responses (Romano et al., 2025).

Despite advances in classification and mechanistic understanding, diagnostic approaches remain limited. Initial evaluation relies primarily on clinical history; however, accumulating evidence indicates that history alone lacks sufficient reliability and diagnostic accuracy. For example, a history of cutaneous reactions to two or more chemically unrelated NSAIDs supports CR diagnosis in NECD patients (Kowalski et al., 2013), whereas oral drug provocation with the suspected NSAID remains the gold standard for confirming NERD(Kowalski et al., 2019). Basophil activation tests have demonstrated suboptimal specificity for NIUAA (Ariza et al., 2014) and oral drug challenge continues to be the reference method for diagnosis. In vitro assays are not widely available, and their predictive value remains uncertain (Ansotegui et al., 2020; Mayorga et al., 2016). Importantly, drug provocation tests carry significant risk and are contraindicated in patients with severe HRs (Barbaud et al., 2024). These limitations underscore the urgent need for reliable biomarkers to improve diagnostic accuracy and risk stratification.

Recent collaborative investigations suggest that COX inhibition alone does not fully explain the phenotypic variability, severity spectrum, or chronic inflammatory features observed in CR phenotypes (Ayuso et al., 2015; Campo et al., 2013; Plaza-Serón et al., 2018; Torres et al., 2021). Emerging evidence implicates oxidative stress as an additional pathogenic mechanism in drug hypersensitivity (Ayuso et al., 2021). Oxidative stress, defined as an imbalance between reactive oxygen species (ROS) production and clearance, profoundly influences inflammatory processes, protein expression, and post-translational modifications (Lei et al., 2015). In NERD, impaired lipoxin biosynthesis has been implicated, reducing anti-inflammatory activity and failing to counterbalance leukotriene-mediated effects, including superoxide generation (Bonnans and Levy, 2007; Kupczyk et al., 2009). Consequently, disruption of the lipoxin– leukotriene equilibrium represents a mechanistic link between redox imbalance and chronic inflammation (Kacprzak and Pawliczak, 2015). Reactive molecules such as peroxynitrite can activate or inhibit COX enzymes depending on substrate availability (Bresson et al., 2012), while oxidation of polyunsaturated fatty acids yields pro-inflammatory mediators such as reactive aldehydes and F2-isoprostanes (Lei et al., 2015). ROS also induce oxidative modifications of cysteine, tyrosine, and methionine residues, which are reversed by thioredoxin and glutathione systems (Mitchell and Marletta, 2005). Furthermore, key inflammatory pathways—including AMPK, NF-κB, and JAK-STAT—are modulated by redox-sensitive changes (Park et al., 2012), while transcription factors such as NF-κB and Nrf2 regulate antioxidant defense, underscoring the bidirectional interplay between oxidative status and inflammation (Morgan and Liu, 2010). Importantly, an association between oxidative stress and drug hypersensitivity has been demonstrated, suggesting that alterations in redox homeostasis may contribute to both CR and SR phenotypes (Ayuso et al., 2021).

The mechanism underlying selective reactions (SRs) to NSAIDs involves activation of the adaptive immune system. Due to their low molecular weight, NSAIDs cannot be directly recognized by immune receptors, necessitating covalent binding to carrier proteins to form hapten–protein complexes that elicit a specific allergic response (Canto et al., 2009). Notably, both parent compounds and reactive metabolites can participate in haptenation by binding covalently to proteins, thereby acting as immunogenic determinants (Ariza et al., 2016). Furthermore, factors that amplify inflammation or generate danger signals may contribute to the development of drug hypersensitivity reactions (DHRs) through mechanisms that remain incompletely understood (Uetrecht and Naisbitt, 2013a). Drug bioactivation can produce highly reactive molecular species, and various biotransformation pathways yield metabolites capable of hapten formation, enabling their conjugation with endogenous proteins and subsequent initiation of an adaptive immune response (Uetrecht and Naisbitt, 2013b). Importantly, an association between oxidative stress and drug hypersensitivity has been demonstrated, suggesting that alterations in redox homeostasis may play a contributory role in the pathogenesis of SRs to NSAIDs.

Genetic predisposition to NSAID-induced hypersensitivity reactions (HRs) remains a central focus of ongoing research; however, the precise contribution of genetic variation to their pathogenesis is still largely undefined. Emerging evidence suggests that genes involved in redox homeostasis may play a critical role, with variants in GSTM1, TXNRD1, and superoxide dismutase (SOD) family genes identified as prominent candidate biomarkers for NSAID-related HRs (Ayuso et al., 2021). To explore genetic determinants that may confer susceptibility to NSAID-induced hypersensitivity reactions and to gain deeper insight into the mechanisms underlying these responses, we conducted a comprehensive analysis of genetic variants in genes implicated in redox homeostasis. Furthermore, we assessed the functional impact of the most salient genetic associations to elucidate their potential biological relevance.

## 2. Results

The detailed characteristics of the CR NSAIDs and SNIUAA patients; as well as the healthy controls with tolerance to NSAIDs are summarized in Table 1 for the discovery phase and replication analysis. The mean age of patients with CRs to NSAIDs and SNIUAA differed significantly compared with NSAIDs tolerant individuals, whereas no statistically significant differences in sex were observed between the two groups in either the discovery phase or the replication cohorts. Ibuprofen and acetylsalicylic acid were the most frequently implicated drugs in CR NSAID patients, whereas metamizole and ibuprofen were the most common culprit drugs in SNIUAA.

**Table 1.**
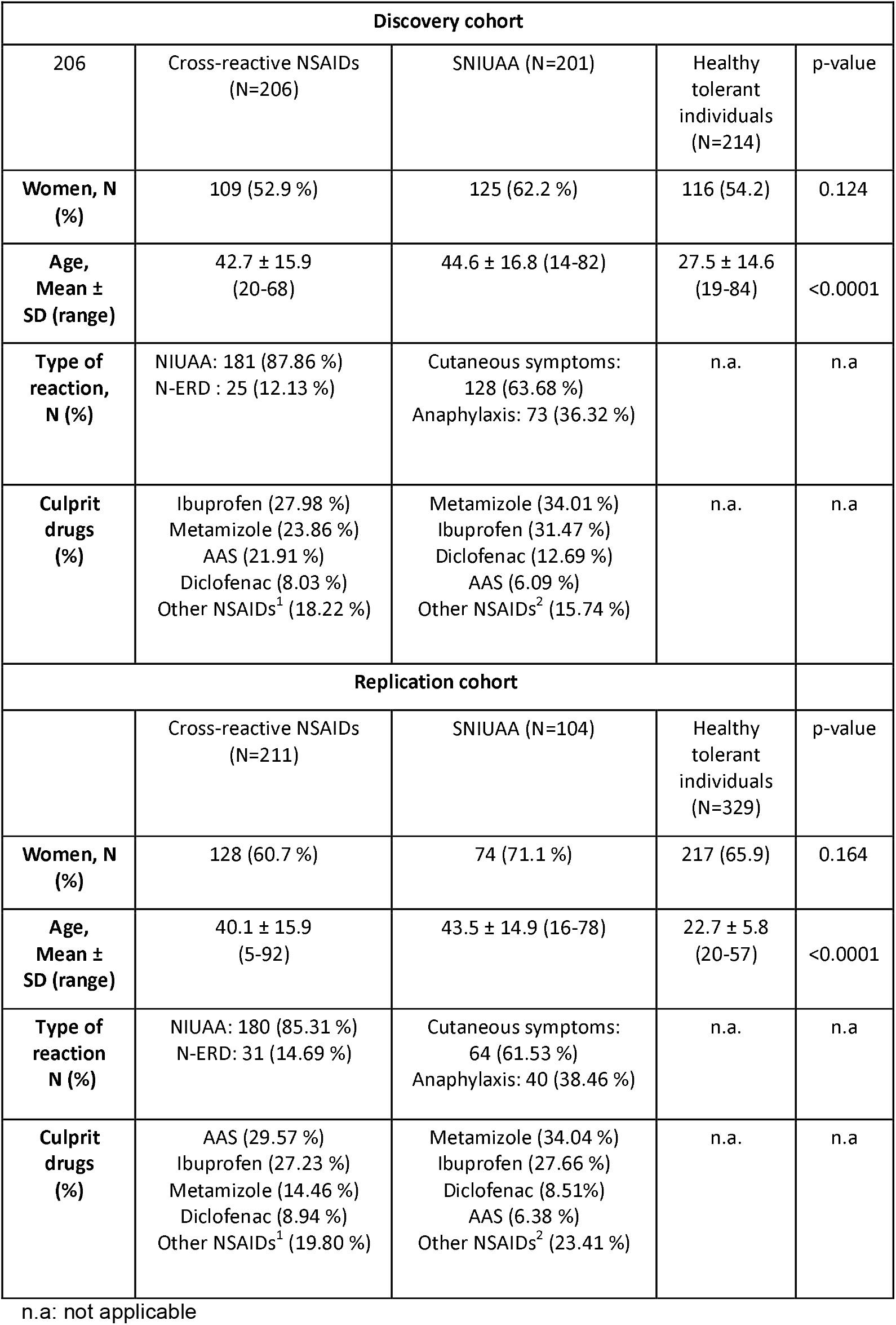
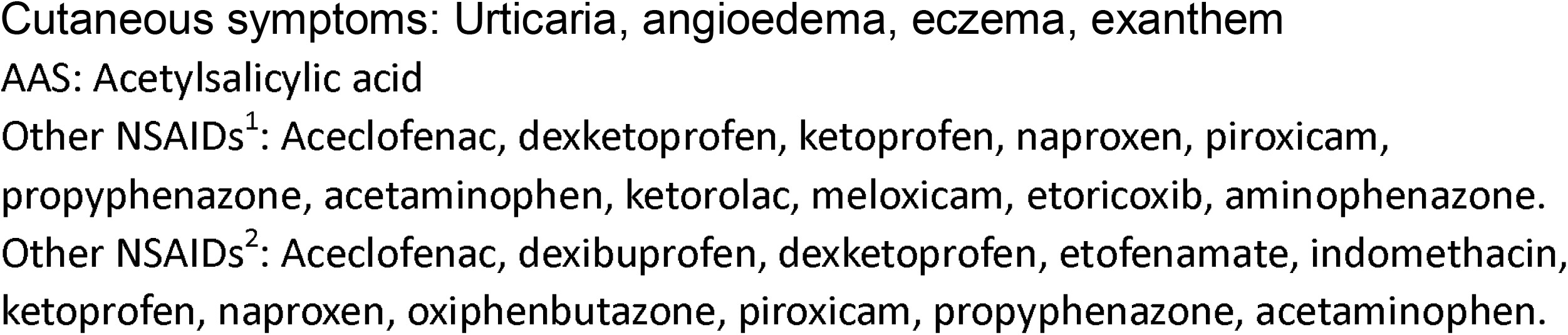
Characteristics of the study population.

### 2.1 Identification of genetic variants associated with cross-reactive NSAIDs hypersensitivity reactions

First, TaqMan genotyping assays were employed to analyze fifty-nine SNVs in the genes described in Table S1. These analyses were performed in a subset of 407 patients along with 214 healthy NSAID-tolerant controls as described in Table 1.

The results of the binary logistic regression analysis are presented in Table 2. Across the evaluated genetic models, three SNVs demonstrated statistically significant associations: rs11551177 in *AKR1C3*, rs1007888 in *GSTT1*, and rs3024678 in *IL4R* (Table 2). Carriers of the *AKR1C3* rs11551177 variant exhibited an increased risk of CR-NSAIDs (P = 0.030; OR = 1.88), as did carriers of the *GSTT1* rs1007888 variant (P = 0.001; OR = 1.91). Conversely, individuals carrying the *IL4R* rs3024678 variant showed a reduced risk of CR-NSAIDs (P = 0.029; OR = 0.029).

**Table 2.**
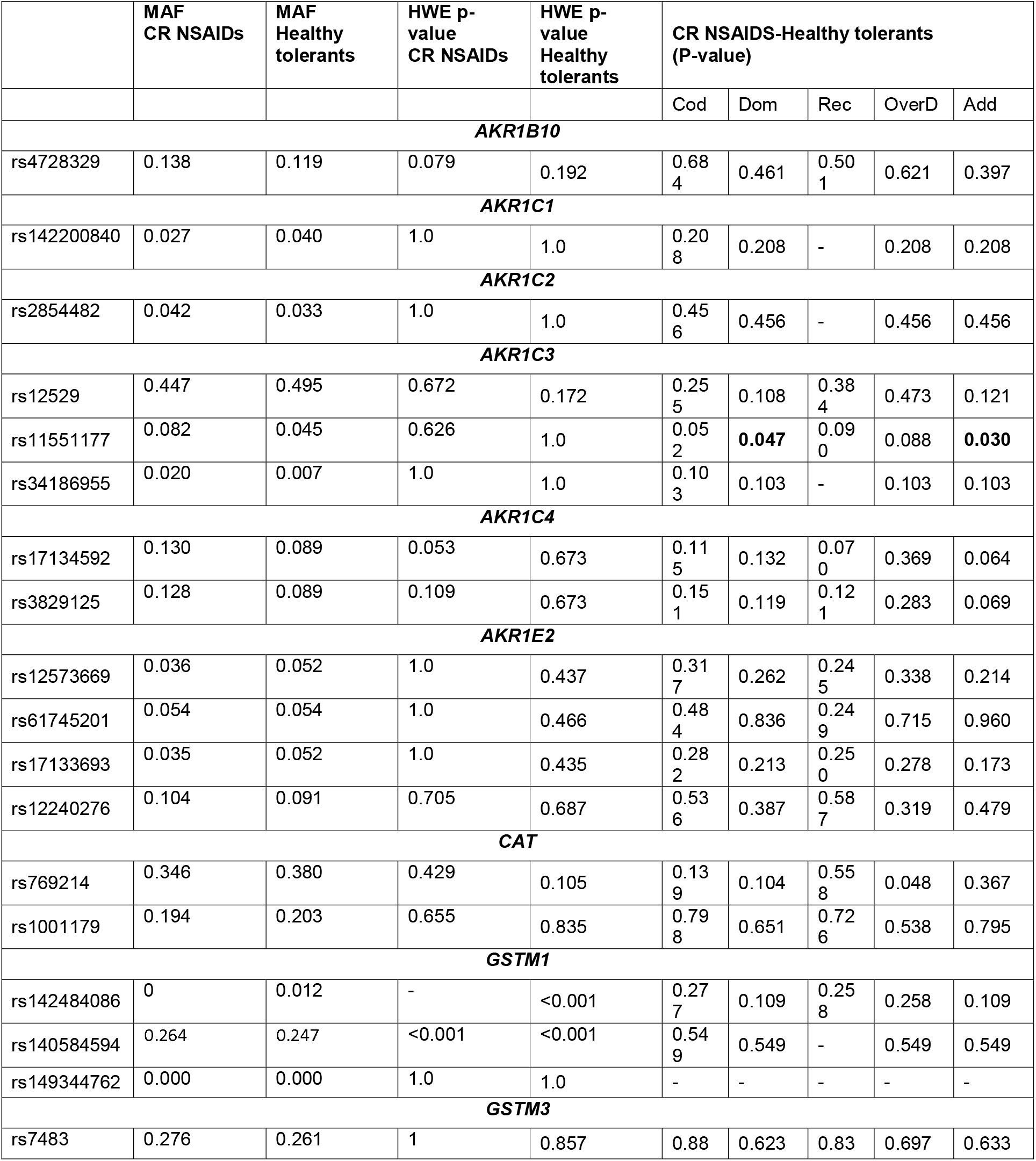
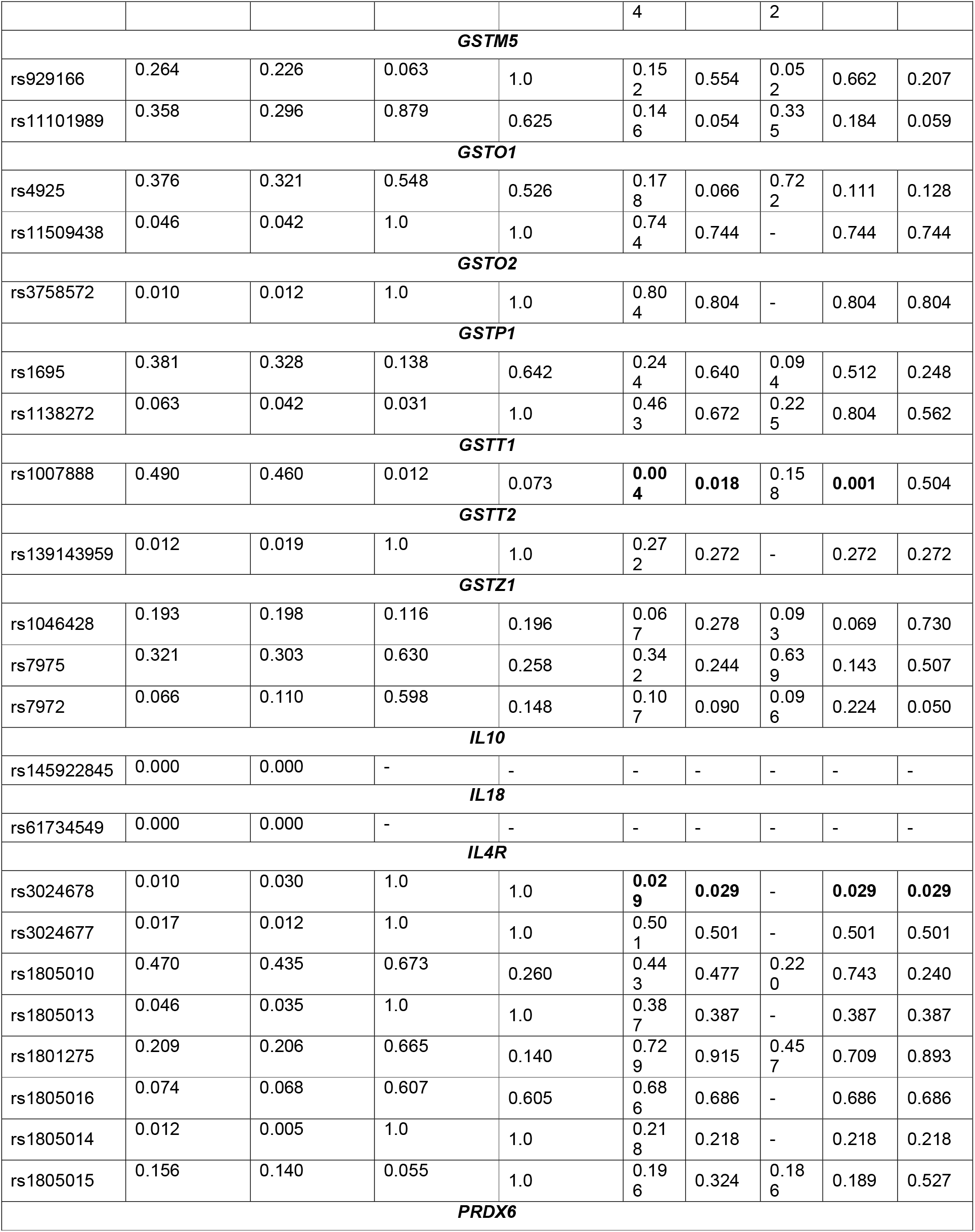
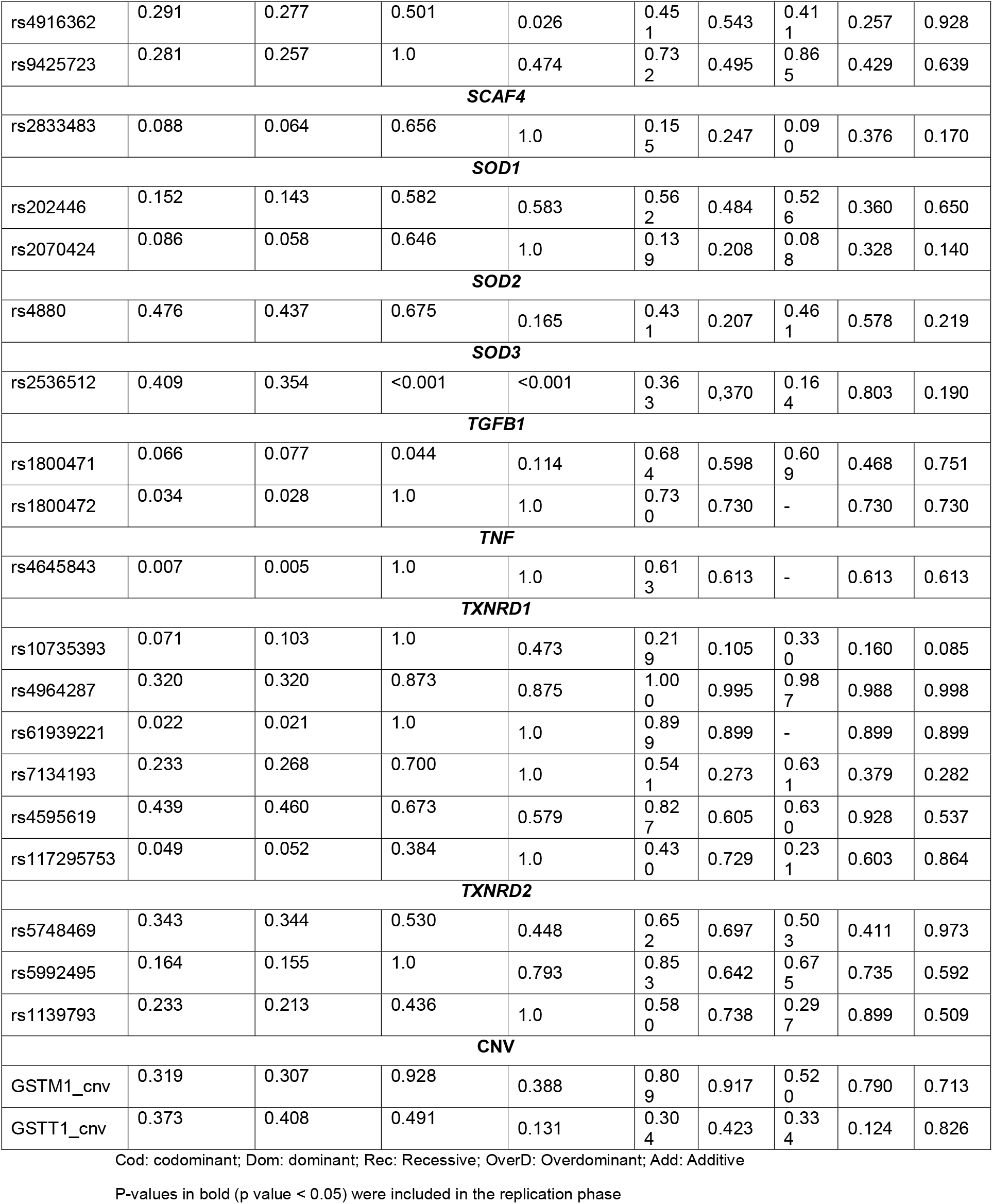
P-values obtained from the genetic association study of CR-NSAIDs at the first stage of the association study.

We also evaluated the potential effects of haplotypes in the genes where two or more SNVs were included in the study: *AKR1C3, AKR1C4, AKR1E2, CAT, GSTM1, GSTM5, GSTO1, GSTP1, GSTT2, GSTZ1, IL4R, PRDX6, SOD1, TGFB1, TXNRD1* and *TXNRD2* genes. A total of 1,349 haplotypes with a frequency greater than 1% were identified in patients from the CR NSAIDs cohort. The haplotypes *TXNRD1* (rs10735393-T, rs7134193-A, rs4595619-A, rs117295753-G; *P-value*: 0.017, OR: 1.86), *AKR1C3* (rs12529-C, rs11551177-G; *P-value*: 0.031, OR: 2.05), *GSTZ1* (rs1046428-C, rs7975-A, rs7972-G; *P-value*: 0.019, OR: 1.47) were associated with an increased risk of developing CR NSAIDs. In contrast, the *IL4R* haplotype (rs3024678-T, rs1805016-T, rs1805015-T; *P-value*: 0.031, OR: 0.22) was associated with a reduced risk of CR NSAIDs. Additionally, we sought to predict the CR NSAIDs risk based on combinations of SNVs by applying logistic regression to model complex genetic interactions, using the logicFS R package (Holger Schwender and Tobias Tietz, 2020). The genotype combination *AKR1E2* rs12573669 TT + *GSTZ1* rs7975 GA/AA + *GSTT1* rs1007888 TC/CC + *AKR1C4* rs17134592 CC/CG was significantly associated with an increased risk of CR NSAIDs (*P-value* < 0.0001, OR: 2.99). This suggests that reduced capacity to neutralize reactive oxygen species (ROS) and electrophiles are related to an increased risk of developing CR NSAID. Conversely, the combination of *SOD1* rs202446 GG + *GSTM5* rs11101989 AA + *TXNRD1* rs4595619 GG/GA-*GSTM1* cnv was associated with a reduced risk of CR NSAIDs (*P-value* < 0.0001, OR: 039).

#### 2.1.1 Replication analyses in patients with cross-reactive NSAIDs hypersensitivity reactions

We included a total cohort of 540 individuals, comprising 211 CR NSAIDs patients and 329 healthy control subjects with no history of DHRs and with confirmed tolerance to NSAIDs (Table 1). Following the initial identification phase, the SNVs analysed at this stage were selected based on statistically significant associations (*P-value* < 0.05) identified through univariate logistic regression and / or haplotype analyses. In addition, SNVs comprising the most statistically significant genetic interaction model were included in the analysis. Consequently, a total of 17 SNVs within the *AKR1C3, AKR1C4, AKR1E2, GSTM5, GSTT1, GSTZ1, IL4R, SOD1* and *TXNRD1* genes were selected for this phase (Table 3).

**Table 3.**
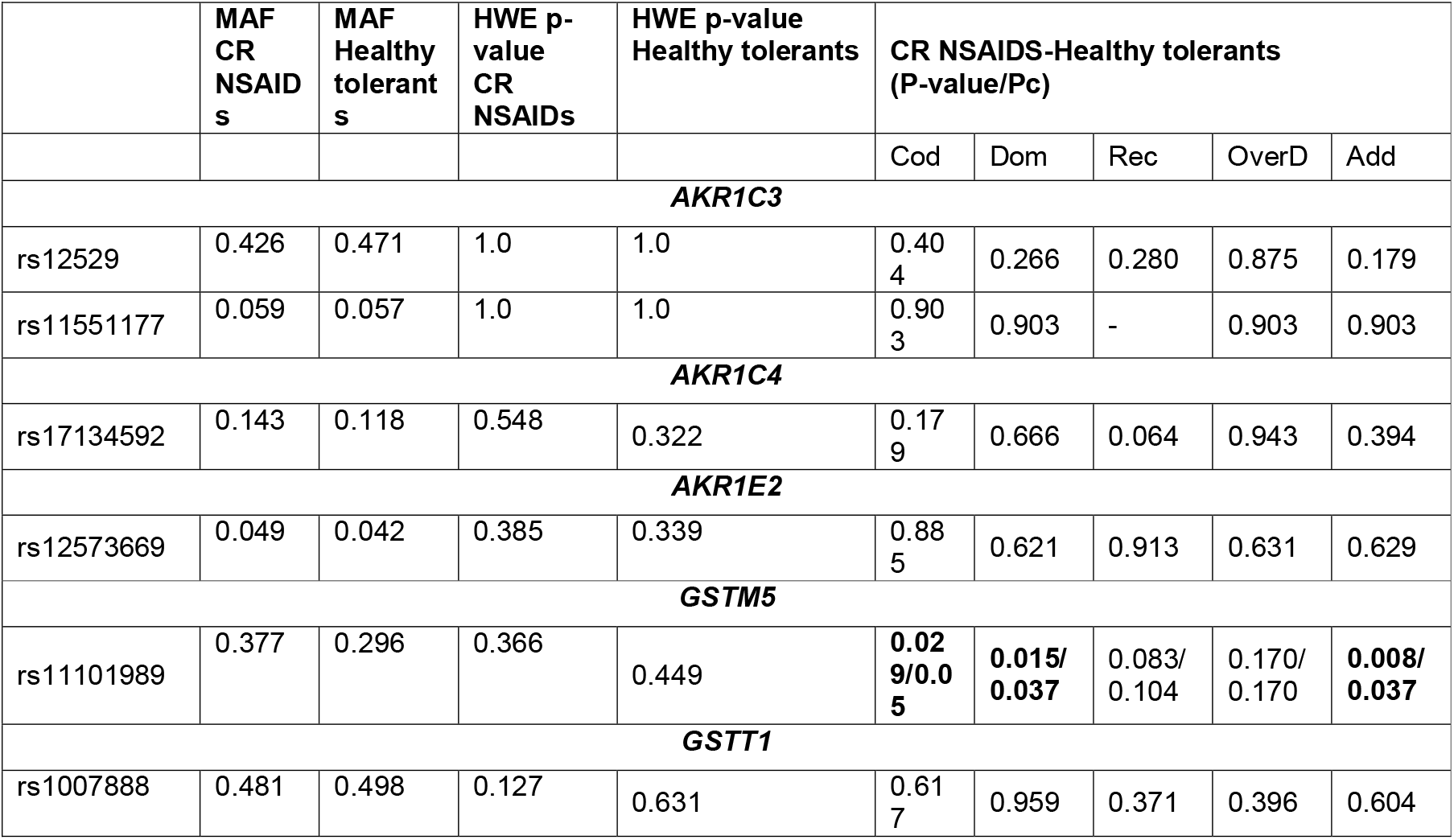
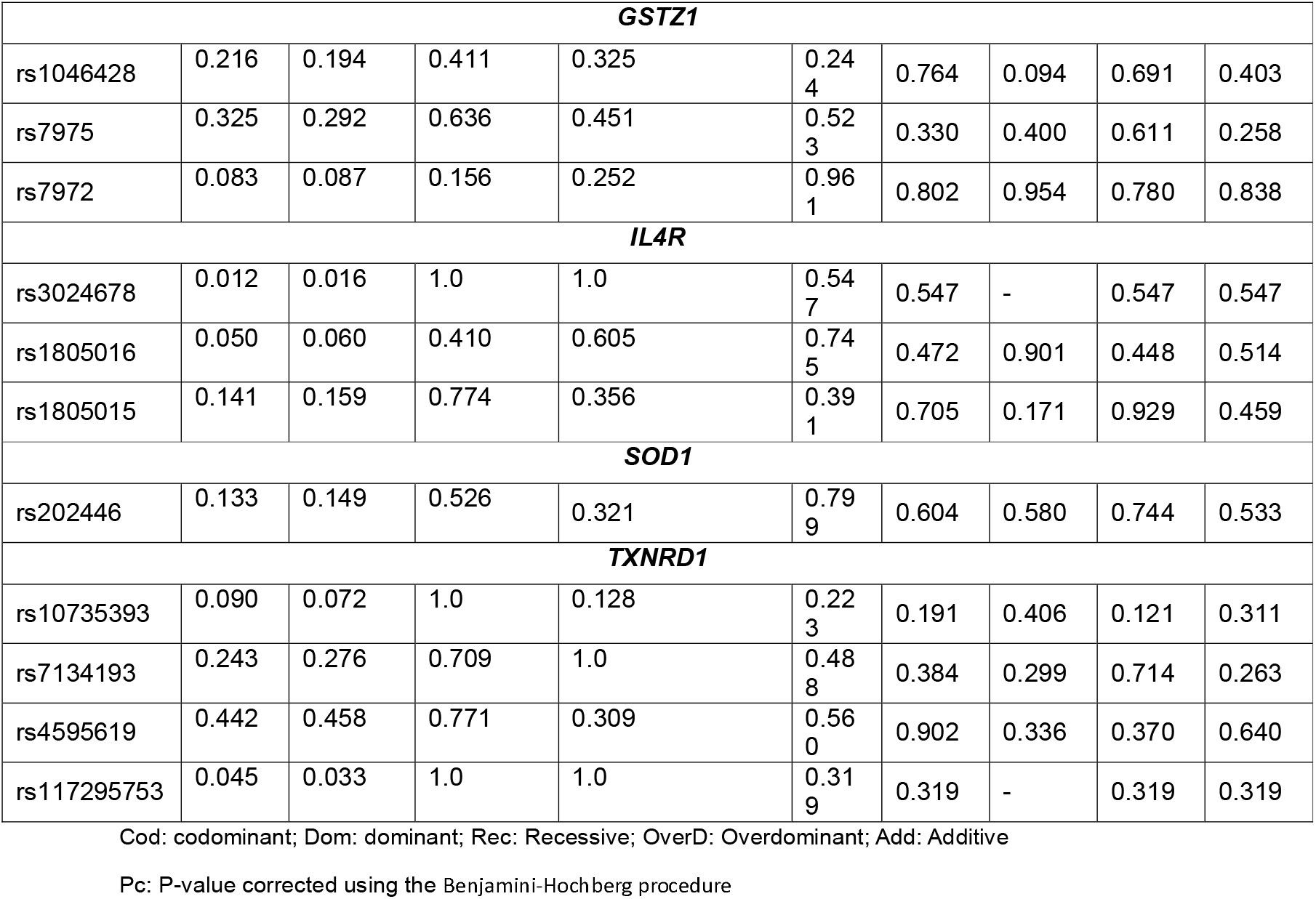
P-values obtained from the genetic association study of CR-NSAIDs at the replication stage of the association study.

The analysis identified a statistically significant association between the SNV *GSTM5* rs11101989 and CR NSAIDs. Individuals carrying the mutant allele of *GSTM5* rs11101989 in homozygosity showed a significantly increased risk of CR NSAIDs (*P-corrected*: 0.037, OR: 1.48) (Table 3). In contrast, the associations previously reported for the SNVs *AKR1C3* rs11551177, *GSTT1* rs1007888 and *IL4R* rs3024678 were not replicated in the validation cohort. Notably, in the discovery study the associations observed for the SNVs *AKR1C3* rs11551177 and *IL4R* rs3024678 were weak, while *GSTT1* rs1007888 did not conform to Hardy-Weinberg equilibrium in the CR NSAIDs cohort (Table 2). To assess the association between these SNVs and susceptibility to CR NSAIDs, the Mantel-Haenszel test was applied to estimate the overall effect and statistical significance combining both cohorts. The results demonstrated that *GSTM5* rs11101989 remained significantly associated with CR NSAIDs (*P-value*: 0.001, OR: 1.40). In contrast, no significant overall associations were observed for *AKR1C3* rs11551177, *GSTT1* rs1007888 and *IL4R* rs3024678 when the discovery and the replication cohorts were combined.

With respect to haplotype analyses, the *GSTZ1* haplotype rs1046428-C + rs7975-A + rs7972-G (*P-value*: 0.010, OR: 1.51) was significantly associated with susceptibility to CR NSAIDs. Replication of these findings in the discovery and validation analyses further supports the role of combined *GSTZ1* variants in modulating the risk of developing CR NSAIDs.

### 2.2 Identification of genetic variants associated with SNIUAA reactions

As previously described, TaqMan genotyping assays were applied to genotype fifty-nine SNVs as shown in Table S1. These analyses were carried out in a defined subset of 201 SNIUAA patients and 214 healthy NSAID-tolerant controls.

The results of the binary logistic regression are summarized in Table 4. Across the different genetic models, ten SNVs exhibited statistically significant associations with SNIUAA. Based on differences in genotype frequencies between groups and the corresponding odd ratios, the variants rs11509438 in *GSTO1*, rs1805016 in *IL4R*, rs12529 in *AKR1C3* and rs7972 in *GSTZ1* were associated with a reduced risk of developing SNIUAA. In contrast, rs1046428 in *GSTZ1*, rs17134592 and rs3829125 in *AKR1C4*, rs2833483 in *SCAF4* and rs2070424 and rs5992495 in *SOD1* and *TXNRD2*, respectively, were associated with an increased risk of developing SNIUAA (Table 4).

**Table 4.**
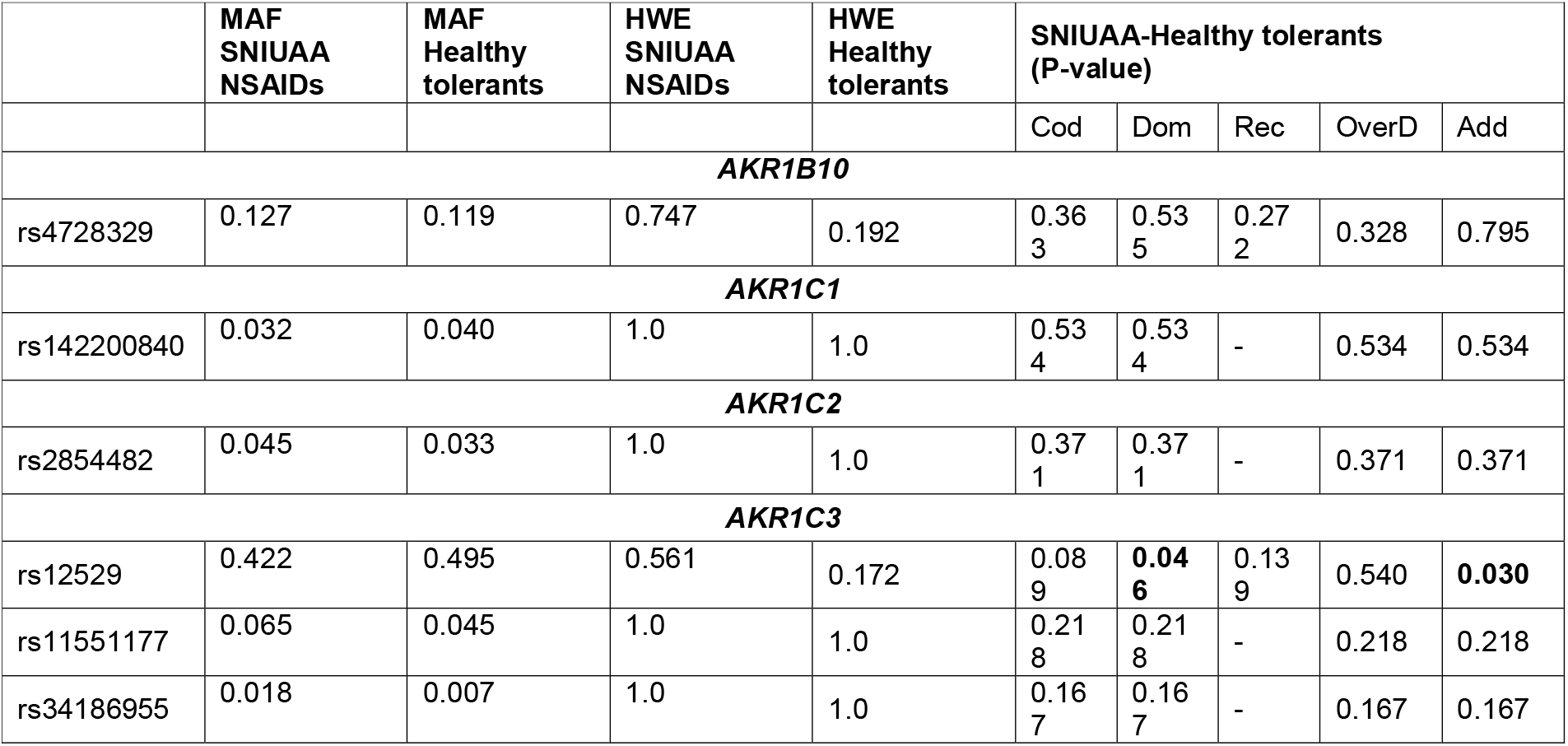
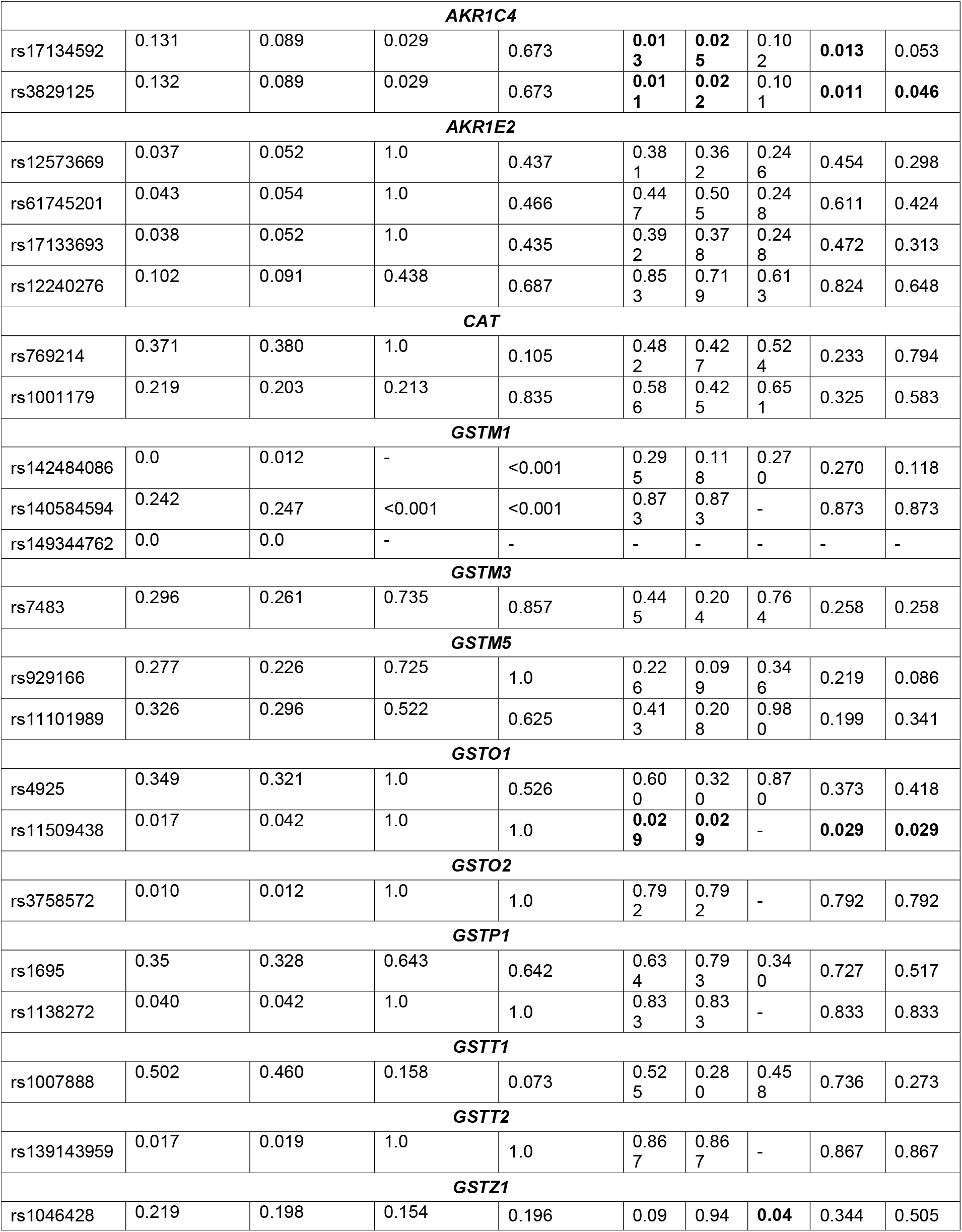
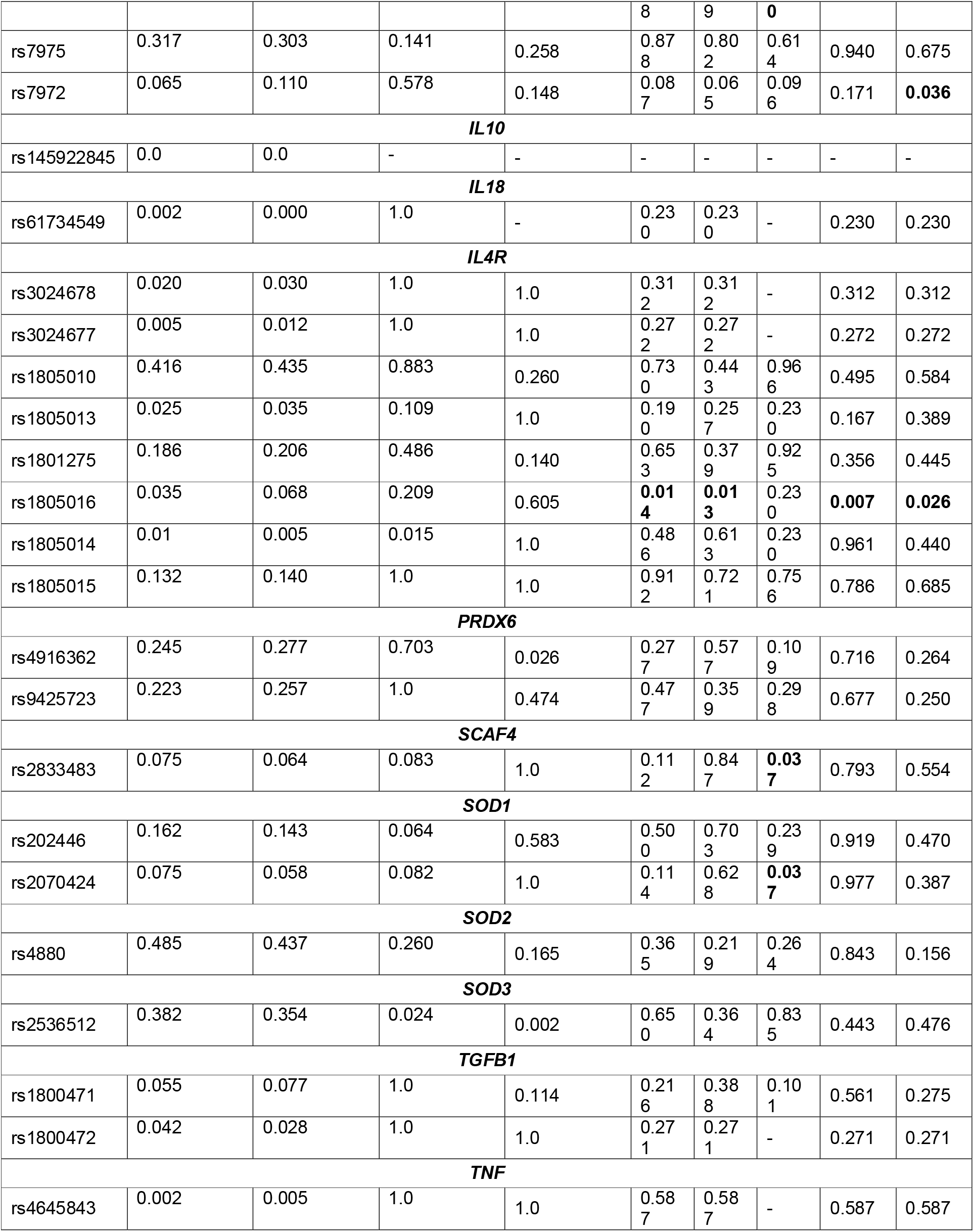
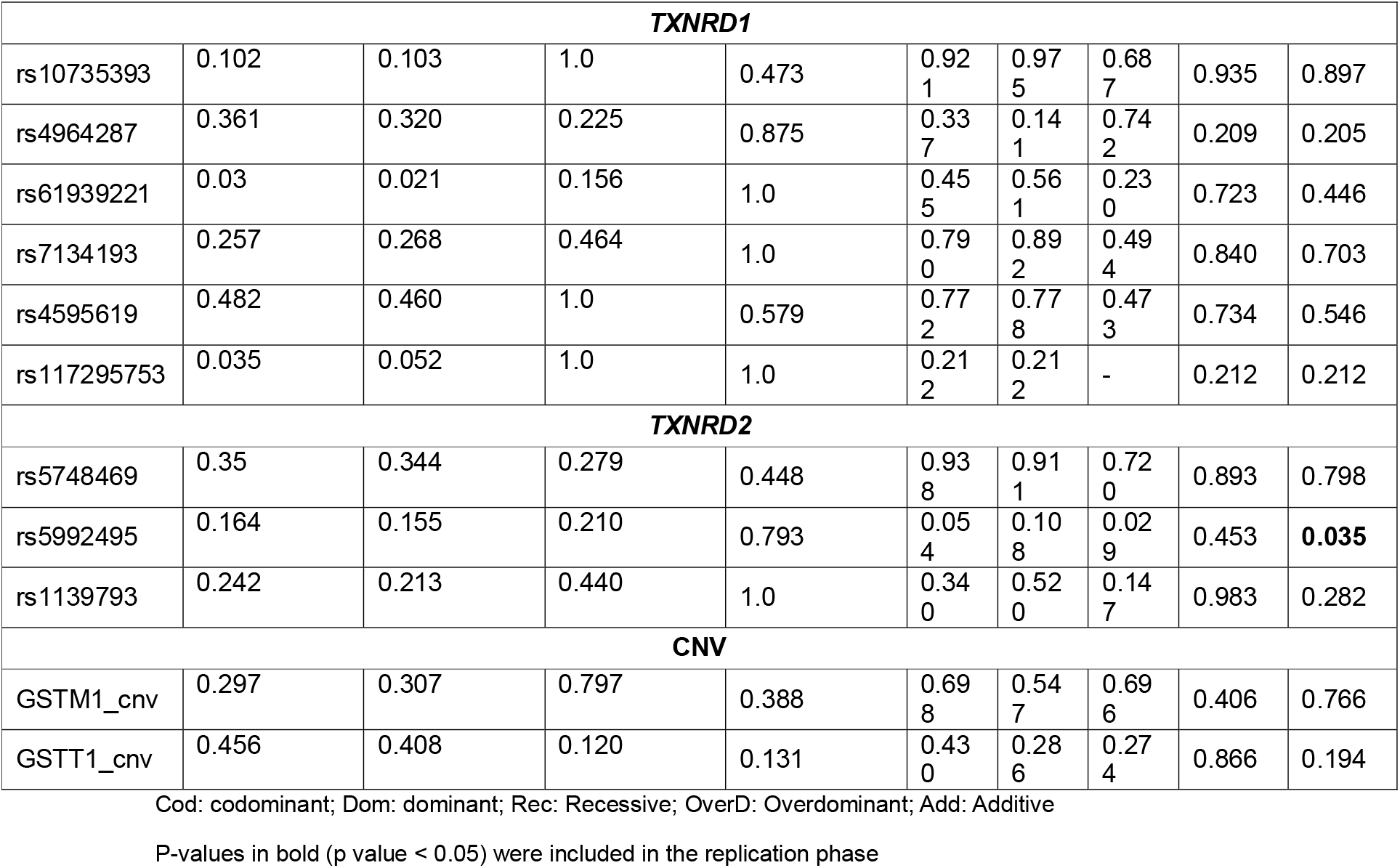
P-values obtained from the genetic association study of SNIUAA at the first stage of the association study.

Haplotype-based association analyses were conducted for the genes where two or more SNVs were included in the study: *AKR1C3, AKR1C4, AKR1E2, CAT, GSTM1, GSTM5, GSTO1, GSTP1, GSTT2, GSTZ1, IL4R, PRDX6, SOD1, TGFB1, TXNRD1* and *TXNRD2* genes. A total of 1,290 haplotypes with frequencies exceeding 1% were identified in the study cohort. The haplotypes *TXNRD1* (rs10735393-T + rs7134193-A + rs4595619-A + rs117295753-G; *P-value*: 0.005, OR: 2.14), *TXNRD2* (rs5992495-G + rs1139793-A; *P-value*: 0.020, OR: 3.29) ) were associated with an increased risk of developing SNIUAA. In contrast, *IL4R* (rs3024678-C + rs1805016-G + rs1805015-T; *P-value*: 0.008, OR: 0.16 ), *AKR1C3* (rs12529-G + rs34186955-C; *P-value*: 0.030, OR: 0.74) and *GSTO1* (rs4925-C + rs11509438-A; *P-value*: 0.032, OR: 0.40) haplotypes ) were associated with a decreased risk of developing SNIUAA.

In addition, logic regression was employed to evaluate whether combinations of SNVs could predict SNIUAA status, allowing the modeling of complex gene-gene interactions using the logicFS R package (Holger Schwender and Tobias Tietz, 2020). A specific multilocus genotype pattern *TXNRD1* rs4964287 CT/TT + *GSTZ1* rs7975 GA/AA + *GSTZ1* rs7972 GG/GA + *PRDX6* rs4916362 AA/AG was significantly associated with an elevated risk of SNIUAA (*P-value* < 0.0001, OR: 2.19). In contrast, the genotype combination comprising *GSTM5* rs929166 TT + *AKR1C2* rs2854482 AA + *AKR1C4* rs3829125 CC + *TXNRD2* rs5992495 TT + *AKR1C3* rs34186955 CC was linked to a decreased risk of SNIUAA (*P-value* < 0.0001, OR: 0.43).

#### 2.2.1 Replication analyses in SNIUAA patients

The study cohort comprised a total of 433 individuals, including 104 patients diagnosed with SNIUAA and 329 control subjects with no history of DHRs and confirmed tolerance to NSAIDs (Table 1). Following the initial discovery phase, the SNVs evaluated at this stage were selected based on statistically significant associations (*P-value* < 0.05) identified through univariate logistic regression and haplotype analysis. Additionally, SNVs consistent with the previously described genetic interaction model were included. Consequently, a total of 12 SNVs across *AKR1C2, AKR1C3, AKR1C4, GSTM5, GSTO1, PRDX6, SOD1, TXNRD1* and *TXNRD2* genes were selected for analysis in this phase (Table 5).

**Table 5.**
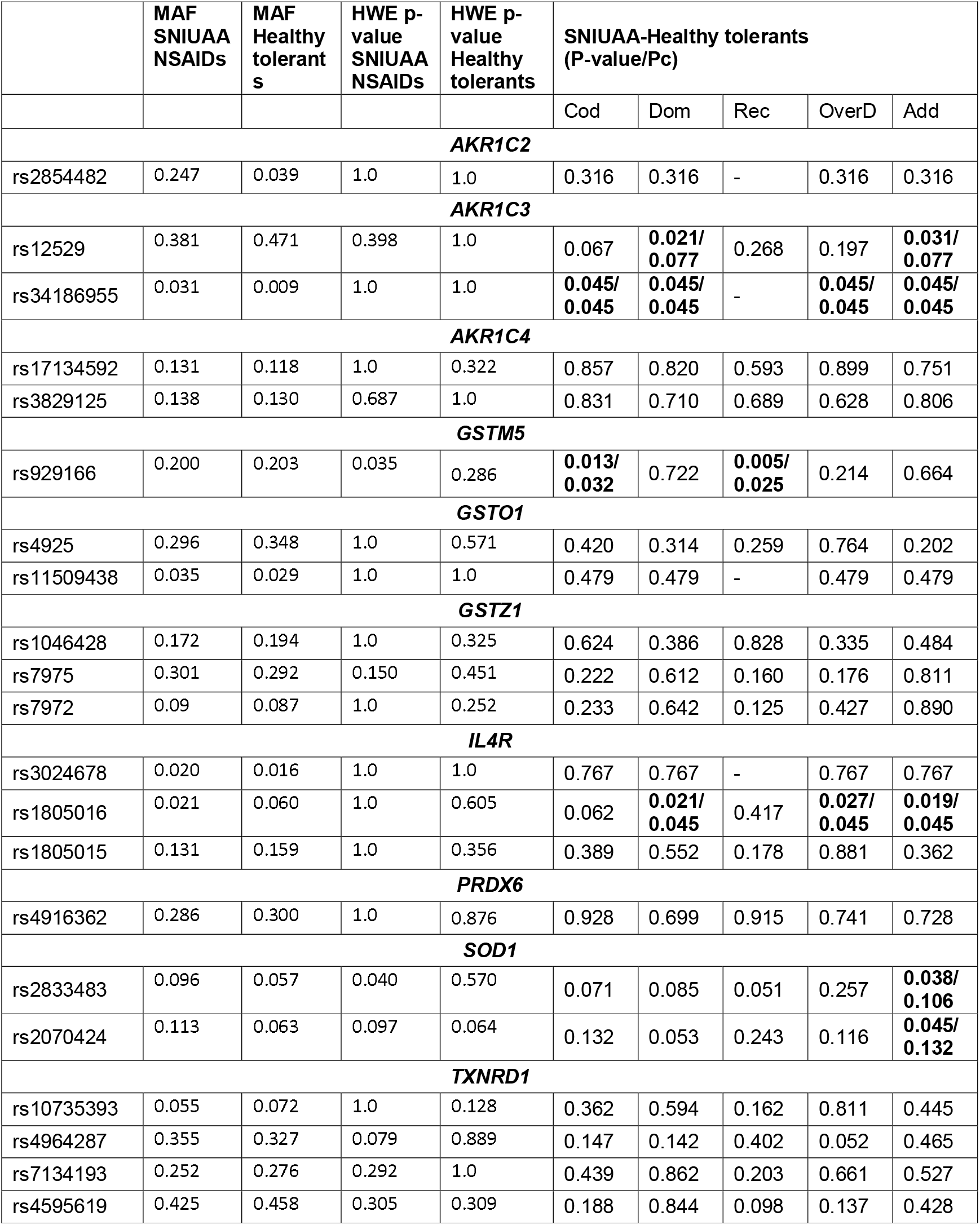
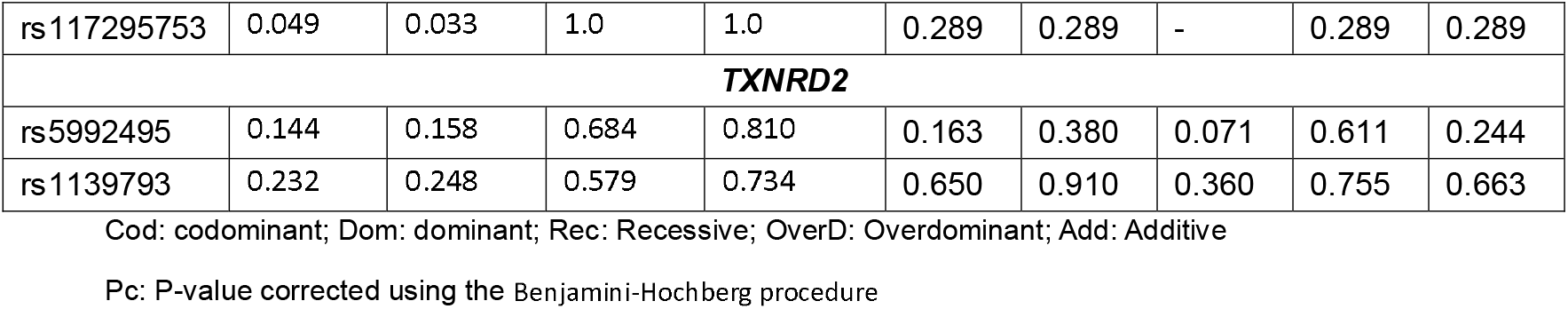
P-values obtained from the genetic association study of SNIUAA at the replication stage of the association study.

|  | MAF<br>SNIUAA<br>NSAIDs | MAF<br>Healthy<br>tolerant<br>s | HWE p-value<br>SNIUAA<br>NSAIDs | HWE p-value<br>Healthy<br>tolerants | SNIUAA-Healthy tolerants<br>(P-value/Pc) |  |  |  |  |
| --- | --- | --- | --- | --- | --- | --- | --- | --- | --- |
|  |  |  |  |  | Cod | Dom | Rec | OverD | Add |
| **AKR1C2** |  |  |  |  |  |  |  |  |  |
| rs2854482 | 0.247 | 0.039 | 1.0 | 1.0 | 0.316 | 0.316 | - | 0.316 | 0.316 |
| **AKR1C3** |  |  |  |  |  |  |  |  |  |
| rs12529 | 0.381 | 0.471 | 0.398 | 1.0 | 0.067 | **0.021/**<br>**0.077** | 0.268 | 0.197 | **0.031/**<br>**0.077** |
| rs34186955 | 0.031 | 0.009 | 1.0 | 1.0 | **0.045/**<br>**0.045** | **0.045/**<br>**0.045** | - | **0.045/**<br>**0.045** | **0.045/**<br>**0.045** |
| **AKR1C4** |  |  |  |  |  |  |  |  |  |
| rs17134592 | 0.131 | 0.118 | 1.0 | 0.322 | 0.857 | 0.820 | 0.593 | 0.899 | 0.751 |
| rs3829125 | 0.138 | 0.130 | 0.687 | 1.0 | 0.831 | 0.710 | 0.689 | 0.628 | 0.806 |
| **GSTM5** |  |  |  |  |  |  |  |  |  |
| rs929166 | 0.200 | 0.203 | 0.035 | 0.286 | **0.013/**<br>**0.032** | 0.722 | **0.005/**<br>**0.025** | 0.214 | 0.664 |
| **GSTO1** |  |  |  |  |  |  |  |  |  |
| rs4925 | 0.296 | 0.348 | 1.0 | 0.571 | 0.420 | 0.314 | 0.259 | 0.764 | 0.202 |
| rs11509438 | 0.035 | 0.029 | 1.0 | 1.0 | 0.479 | 0.479 | - | 0.479 | 0.479 |
| **GSTZ1** |  |  |  |  |  |  |  |  |  |
| rs1046428 | 0.172 | 0.194 | 1.0 | 0.325 | 0.624 | 0.386 | 0.828 | 0.335 | 0.484 |
| rs7975 | 0.301 | 0.292 | 0.150 | 0.451 | 0.222 | 0.612 | 0.160 | 0.176 | 0.811 |
| rs7972 | 0.09 | 0.087 | 1.0 | 0.252 | 0.233 | 0.642 | 0.125 | 0.427 | 0.890 |
| **IL4R** |  |  |  |  |  |  |  |  |  |
| rs3024678 | 0.020 | 0.016 | 1.0 | 1.0 | 0.767 | 0.767 | - | 0.767 | 0.767 |
| rs1805016 | 0.021 | 0.060 | 1.0 | 0.605 | 0.062 | **0.021/**<br>**0.045** | 0.417 | **0.027/**<br>**0.045** | **0.019/**<br>**0.045** |
| rs1805015 | 0.131 | 0.159 | 1.0 | 0.356 | 0.389 | 0.552 | 0.178 | 0.881 | 0.362 |
| **PRDX6** |  |  |  |  |  |  |  |  |  |
| rs4916362 | 0.286 | 0.300 | 1.0 | 0.876 | 0.928 | 0.699 | 0.915 | 0.741 | 0.728 |
| **SOD1** |  |  |  |  |  |  |  |  |  |
| rs2833483 | 0.096 | 0.057 | 0.040 | 0.570 | 0.071 | 0.085 | 0.051 | 0.257 | **0.038/**<br>**0.106** |
| rs2070424 | 0.113 | 0.063 | 0.097 | 0.064 | 0.132 | 0.053 | 0.243 | 0.116 | **0.045/**<br>**0.132** |
| **TXNRD1** |  |  |  |  |  |  |  |  |  |
| rs10735393 | 0.055 | 0.072 | 1.0 | 0.128 | 0.362 | 0.594 | 0.162 | 0.811 | 0.445 |
| rs4964287 | 0.355 | 0.327 | 0.079 | 0.889 | 0.147 | 0.142 | 0.402 | 0.052 | 0.465 |
| rs7134193 | 0.252 | 0.276 | 0.292 | 1.0 | 0.439 | 0.862 | 0.203 | 0.661 | 0.527 |
| rs4595619 | 0.425 | 0.458 | 0.305 | 0.309 | 0.188 | 0.844 | 0.098 | 0.137 | 0.428 |
| rs117295753 | 0.049 | 0.033 | 1.0 | 1.0 | 0.289 | 0.289 | - | 0.289 | 0.289 |
| ***TXNRD2*** |  |  |  |  |  |  |  |  |  |
| rs5992495 | 0.144 | 0.158 | 0.684 | 0.810 | 0.163 | 0.380 | 0.071 | 0.611 | 0.244 |
| rs1139793 | 0.232 | 0.248 | 0.579 | 0.734 | 0.650 | 0.910 | 0.360 | 0.755 | 0.663 |

Cod: codominant; Dom: dominant; Rec: Recessive; OverD: Overdominant; Add: Additive
Pc: P-value corrected using the Benjamini-Hochberg procedure

The analysis revealed several statistically significant associations between SNVs in *AKR1C3, GSTM5, IL4R* and *SOD1* genes and SNIUAA. Thus, carriers of the variant alleles *AKR1C3* rs34186955 (*P-value*: 0.045, OR: 3.69) or *SOD1* rs2070424 rs2833483 (*P-value*: 0.045, OR: 1.74 and *P-value*: 0.038, OR: 1.84; respectively) exhibited a significantly increased risk of SNIUAA (Table 4). In contrast, individuals carrying the variant allele of *IL4R* rs1805016 (*P-value*: 0.019, OR: 0.33), or *GSTM5* rs929166 (*P-value*: 0.005) or *AKR1C3* rs12529 (*P-value*: 0.021, OR: 0.57) showed a decreased risk of developing SNIUAA (Table 5). However, after correction for multiple testing, only *AKR1C3* rs34186955 (*P-corrected*: 0.045), *GSTM5* rs929166 (*P-corrected*: 0.025) and *IL4R* rs1805016 (*P-corrected*: 0.045) remained statistically significant. Conversely, the rest of previously reported associations in the discovery phase were not confirmed in the validation cohort. It should be noted that the SNVs *GSTO1* rs11509438, *GSTZ1* rs1046428 or rs7972 and *TXNRD2* rs5992495 showed only modest associations, whereas *AKR1C4* rs17134592 or rs3829125 deviated from Hardy-Weinberg equilibrium in the SNIUAA cohort (Table 4). To further evaluate the relationship between these variants and the susceptibility to SNIUAA, a Mantel-Haenszel analysis was performed to estimate the pooled effect size and corresponding statistical significance across both cohorts. This combined analysis confirmed that *SOD1* rs2070424 (*P-value*: 0.03, OR: 1.50), rs2833483 (*P-value*: 0.03, OR: 7.71), *IL4R* rs1805016 (*P-value*: 0.001, OR: 0.38), *AKR1C3* rs12529 (*P-value*: 0.002, OR: 0.61) and *AKR1C3* rs34186955 (*P-value*: 0.02, OR: 3.08) remained significantly associated with SNIUAA.

Finally, haplotype analysis demonstrated that the *AKR1C3* haplotype defined by rs12529-G + rs34186955-C was significantly associated with a reduced risk of SNIUAA (*P-value*: 0.017, OR: 0.57). In contrast, previously reported haplotypes associations involving *TXNRD1, TXNRD2, IL4R* and *GSTO1* were not replicated in the validation cohort.

### 2.3 Functional validation

The genetic association analysis indicated that carriers of the variant allele of *GSTM5* rs11101989 had a significantly higher susceptibility to CR NSAIDs. This SNV is located in a non-coding region at the 5′region of the *GSTM5* gene. Based on this finding, we investigated whether this SNV influences serum GST activity in patients with CR NSAIDs. Serum GST activity was assessed in 22 patients with the rs11101989 AA genotype, 22 patients with the AC genotype, and 8 patients with the CC genotype. A statistically significant difference in GST activity was observed across genotypes (*P-value*: 0.003), with patients carrying the rs11101989 CC genotype exhibiting the lowest mean serum GST activity (mean: 2.81 nmol/min/ml; SEM: 1.18), that is, about a three-fold decrease in GSTM5 enzyme activity as compared with carriers of non-mutated (rs11101989 AA or AC genes; (Figure 2), thus providing a functional basis for the association identified in the replication cohort of CR patients. Serum GST activity was also assessed with regard to the *GSTZ1* haplotype that were identified as a risk factor for CR NSAIDS. For this, serum from 21 individuals lacking the *GSTM5* rs11101989 CC genotype was examined. No statistically significant association was detected between serum GST activity and the analyzed *GSTZ1* haplotype (Figure 3).

**Figure 1.**
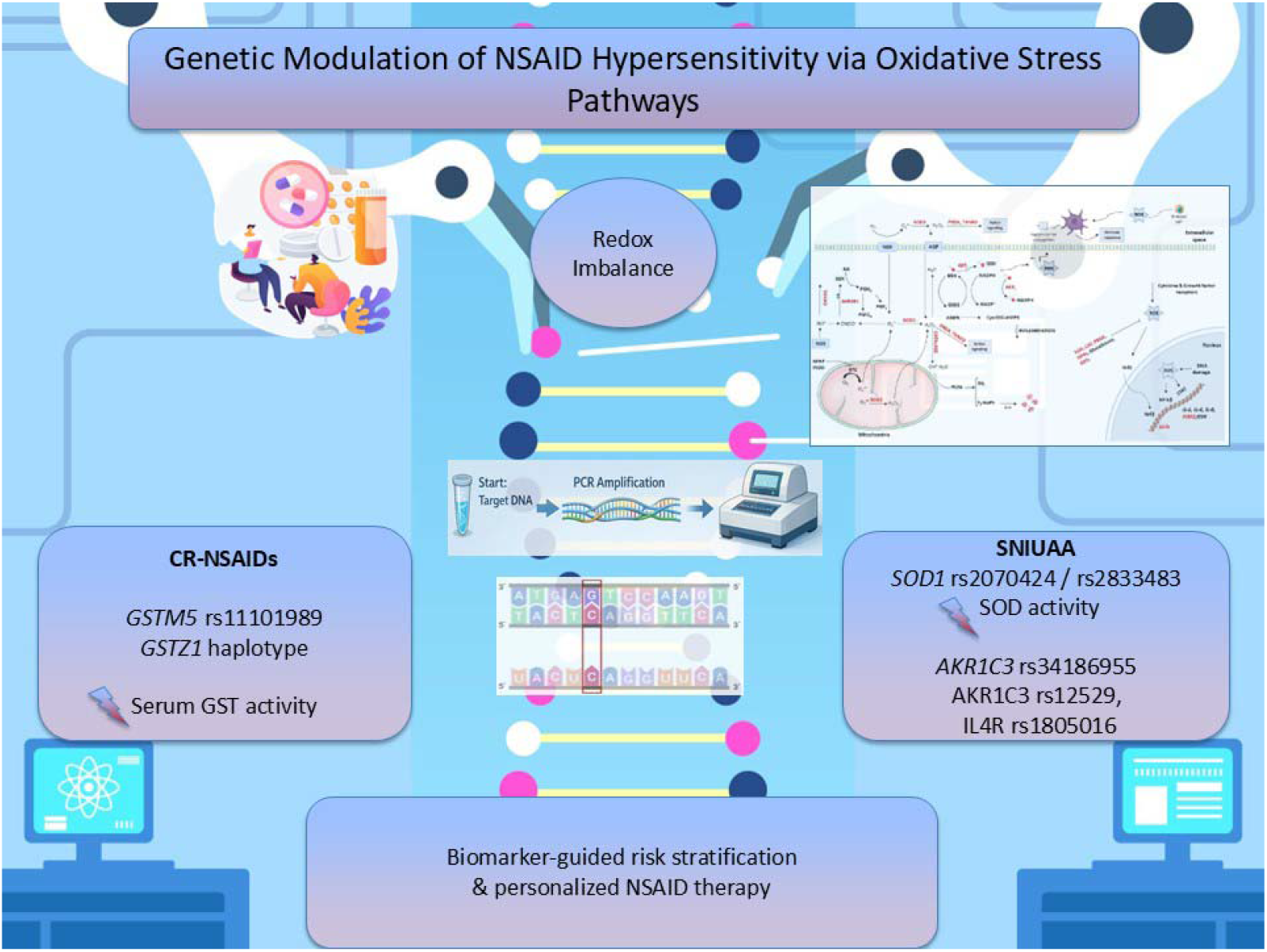
Graphical abstract

**Figure 2.**
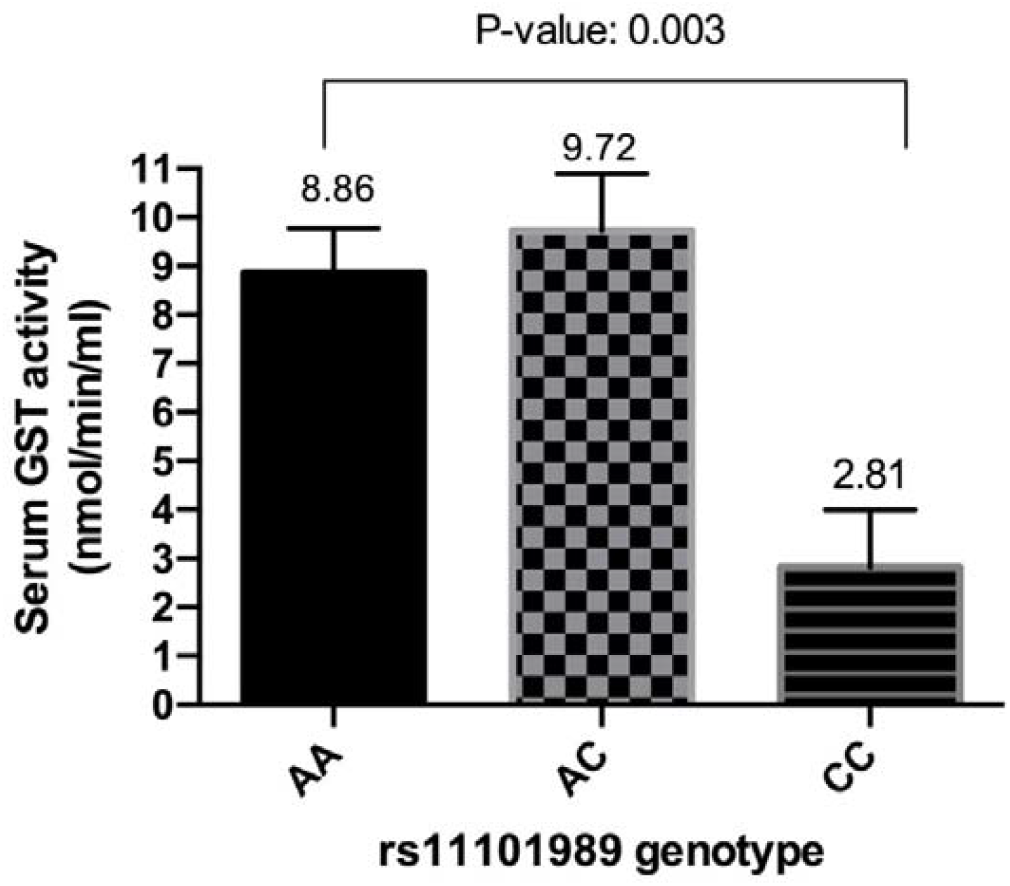
Effect of *GSTM5* rs11101989 genotype on the serum GST activity (mean ± standard error of mean) in CR NSAIDs patients

**Figure 3.**
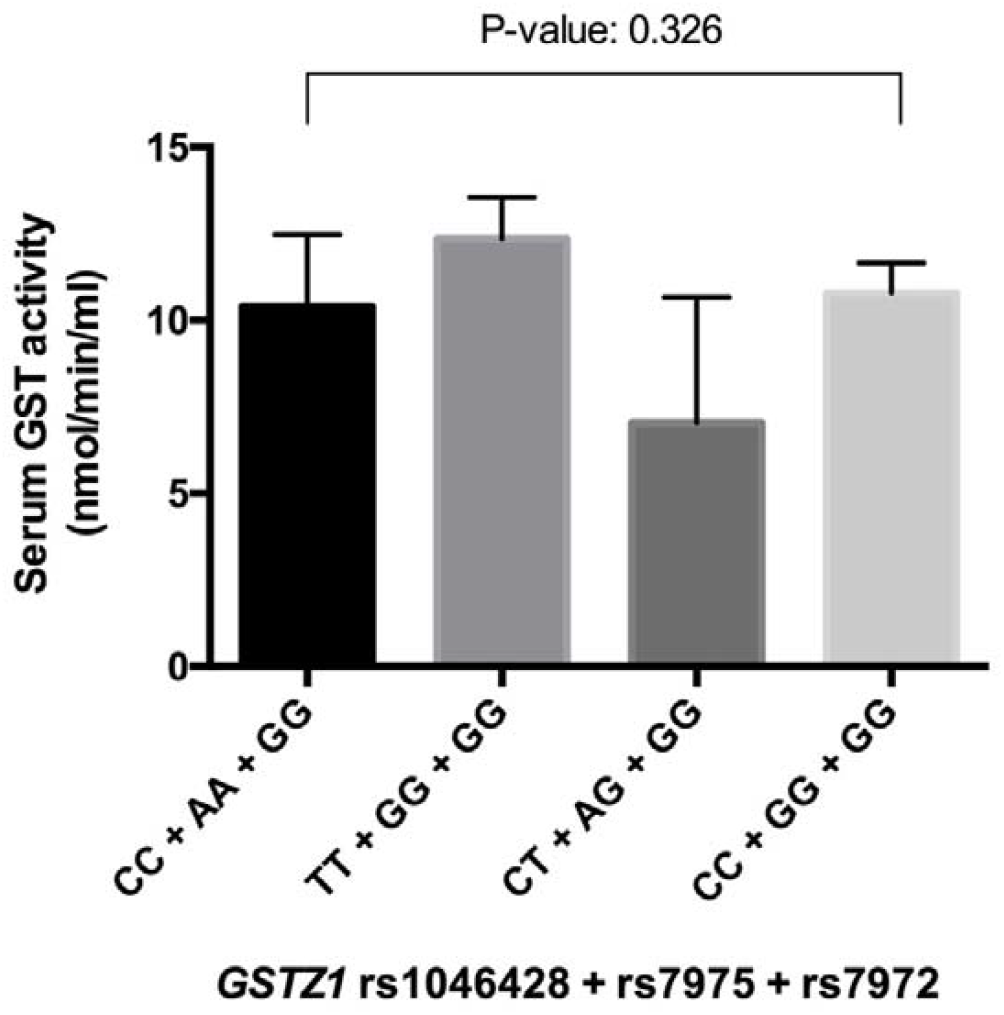
Effect of *GSTZ1* rs1046428 rs7975 rs7972 haplotypes on the serum GST activity (mean ± standard error of mean) in CR NSAIDs patients

Based on the findings of the genetic association analysis in the SNIUAA patient cohort, the intronic variant *SOD1* rs2070424 was associated with a significantly increased risk of SNIUAA both in the discovery and the replication cohorts. Considering this result, we examined whether this gene variant affects SOD activity. Serum SOD activity was measured in 15 patients carrying the rs2070424 AA genotype, 20 patients with the AG genotype, and 5 patients with the GG genotype. A statistically significant difference in SOD activity was detected among the genotypic groups (*P-value*: 0.003), with patients carrying the GG genotype showing the lowest mean serum SOD activity (mean: 0.14 U/ml; SEM: 0.09) (Figure 4). Therefore, our findings indicate that a genetic variant of the SOD enzyme that causes decreased enzyme activity, is associated with increased risk of developing SNIUAA.

**Figure 4.**
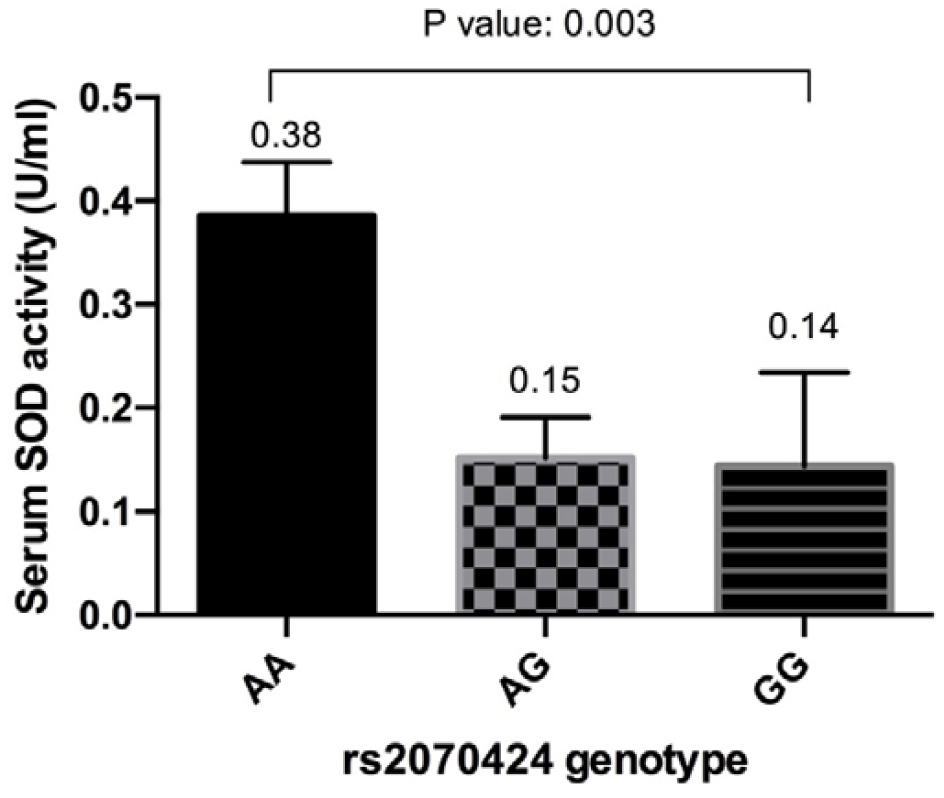
Effect of *SOD1* rs2070424 genotype on the serum SOD activity (mean ± standard error of mean) in SNIUAA patients

In addition, the nonsynonymous variants *AKR1C3* rs34186955 and rs12529, as well as the haplotype formed by these variants, were significantly associated with SNIUAA risk. Based on these results, we investigated whether these variants or their corresponding haplotype affect serum AKR activity. Thus, AKR activity was measured in 27 patients carrying the rs34186955 CC genotype (mean: 10.11 µmol/min/ml; SEM: 0.09) and 7 patients with the CT genotype (mean: 10.72 µmol/min/ml; SEM: 0.72) (Figure S2). Additionally, AKR activity was also measured in 11 patients carrying the rs12529 CC genotype (mean: 10.46 µmol/min/ml; SEM: 0.46), 11 patients carrying the CG genotype (mean: 10.07 µmol/min/ml; SEM: 0.17) and 9 patients with the GG genotype (mean: 10.19 µmol/min/ml; SEM: 0.11) (Figure S3). Furthermore, the effect of *AKR1C3* haplotype in AKR activity was assessed. Therefore, AKR activity was also measured in 9 patients carrying the *AKR1C3* rs34186955 + rs12529 CC + GG genotype (mean: 10.19 µmol/min/ml; SEM: 0.11), 7 patients carrying the CC + CC genotype (mean: 9.97 µmol/min/ml; SEM: 0.13), 8 patients with the CC + CG genotype (mean: 10.13 µmol/min/ml; SEM: 0.22), 4 patients with the CT + CC genotype (mean: 11.33 µmol/min/ml; SEM: 1.23) and 3 patients with the CT + CG genotype (mean: 9.92 µmol/min/ml; SEM: 0.23) (Figure S4). However, no statistically significant association was observed between serum AKR activity and either the analyzed *AKR1C3* SNVs or the *AKR1C3* haplotype.

## 3 Discussion

In this study, we performed a comprehensive genetic and functional analysis of redox homeostasis–related genes in a large, well-characterized cohort of patients with hypersensitivity reactions (HRs) to NSAIDs. By integrating discovery and replication analyses with serum enzymatic activity measurements, our findings provide novel evidence supporting a contributory role of oxidative stress pathways in both CR NSAIDs and SNIUAA phenotypes. Overall, these results reinforce the concept that redox imbalance represents an additional, mechanistically relevant contributor in the pathogenesis of HRs to NSAIDs.

The most robust and reproducible genetic association was observed for the SNV *GSTM5* rs11101989. This association remained statistically significant after replication and pooled analysis, underscoring *GSTM5* as a relevant susceptibility locus for CR NSAID reactions. Importantly, functional evaluation demonstrated that carriers of the mutant *GSTM5* rs11101989 CC genotype exhibited significantly reduced serum GST activity, providing biological plausibility for the genetic findings (Figure 1). This evidence aligns with previous reports indicating that variability in GST expression may contribute to interindividual differences in susceptibility to DHRs to NSAIDs (den Braver et al., 2016; Lucena et al., 2008; Sánchez-Gómez et al., 2016). *GSTM5* belongs to the GST family and encodes glutathione S-transferase mu 5, which is expressed in intracellular compartments, including mitochondria. This enzyme plays an important role in maintaining cellular redox homeostasis by protecting organelles from oxidative damage. Genetic variation within *GSTM5* has been reported to influence enzymatic function and may alter susceptibility to oxidative stress (Butrym et al., 2021).

The variant rs11101989 is located within the *GSTM* cluster on chromosome 1p13.3, which comprises *GSTM1–GSTM5* genes (Pearson et al., 1993). Expression-based analyses of this genomic region have identified rs11101989 as part of an expression quantitative trait locus (eQTL), with reported associations between this variant and *GSTM5* expression levels in liver tissue (Schadt et al., 2008). In addition, rs11101989 has been linked to disease-free survival in patients with acute myeloid leukemia (Yee et al., 2013). To our knowledge, the present study is the first to investigate the impact of *GSTM5* rs11101989 on serum GST activity, demonstrating an association with the risk of developing CR NSAIDs. GSTs play a critical role in detoxifying reactive oxygen species and electrophilic metabolites (Sánchez-Gómez et al., 2016). Reduced GST activity may lead to increased oxidative stress during NSAID exposure. In the context of COX inhibition, such redox imbalance may amplify inflammatory signaling cascades, thereby increasing the risk for HRs to NSAIDs (Ayuso et al., 2021).

In addition to *GSTM5* rs11101989, haplotype analysis revealed a significant association between *GSTZ1* variants and CR NSAID susceptibility. *GSTZ1* catalyzes the terminal steps of phenylalanine and tyrosine catabolism and contributes to detoxification of xenobiotics relevant in drug metabolism (Stacpoole, 2023). Published evidence supports that haplotype variation in *GSTZ1* defined by the nonsynonymous variants rs1046428, rs7975, and rs7972 influences the metabolism of dichloroacetic acid, suggesting that these SNVs may modulate enzyme activity (Shroads et al., 2012a). However, our functional study revealed that this haplotype did not influence total serum GST activity. Given the dual role of *GSTZ1* in the metabolism of endogenous and exogenous reactive species, it is plausible that *GSTZ1* haplotypes modulate substrate-specific redox processes that are not fully captured by total GST activity assays (Shroads et al., 2012b).

For SNIUAA reactions, different SNVs in redox-related genes showed significant associations, including variants in *SOD1, AKR1C3, GSTM5*, and *IL4R* in the validation cohort. Combined analysis confirmed associations for *SOD1* rs2070424 and rs2833483, *IL4R* rs1805016, and *AKR1C3* rs12529 and rs34186955, as well as the *AKR1C3* haplotype defined by rs12529-G and rs34186955-C. *IL4R* encodes the interleukin-4 receptor, a key mediator of type 2 immune signaling that governs leukocyte activation, proliferation, and differentiation. Beyond its immunological role, IL-4 has also been shown to influence glutathione biosynthesis, thereby linking immune signaling with cellular redox homeostasis (Ryan et al., 2012).

The SNV *IL4R* rs1805016 is a nonsynonymous variant (Ser752Ala) associated with impaired signal transduction and consequent alterations in IL4R functional activity (Forster et al., 2004). Regarding *AKR1C3* variants, *AKR1C3* encodes an enzyme involved in steroid hormone metabolism and prostaglandin signaling downstream of COX activity, influencing inflammatory and proliferative responses (50,51). Certain NSAIDs such as indomethacin, flufenamic acid, diclofenac, or ibuprofen may bind to the AKR1C3 active site and act as competitive or mixed inhibitors, suppressing its prostaglandin-reducing activity through a COX-independent mechanism(72,73). The SNVs rs12529 and rs34186955 results in a missense substitution His5Gln and Pro180Ser; respectively. In case of *AKR1C3* rs12529, the amino acid replaced shares common chemical properties; nonetheless this change alters a putative exonic splicing enhancer motif that may cause alternative splicing regulatory effects (Shaon et al., 2025; Yu et al., 2013). Based on *in vitro* functional characterization, the *AKR1C3* rs34186955 mutant variant exhibited reduced reductase activity compared with the wild-type variant; particularly for prostaglandin substrates (Takano et al., 2023). Although our functional study revealed that these SNVs in *AKR1C3* or the haplotype composed by them did not influence total serum AKR activity in SNIUAA patients.

Notably, the SNVs *SOD1* rs2070424 and rs2833483 were associated with reduced serum SOD activity, suggesting impaired superoxide detoxification that may facilitate oxidative protein modification, hapten formation, and immune activation. *SOD1* catalyzes the conversion of superoxide radicals into hydrogen peroxide and molecular oxygen, exerting a central role in intracellular redox homeostasis (Ighodaro and Akinloye, 2018). The SNVs *SOD1* rs2070424 and rs2833483 are non-coding variants located within the intronic region and the 3′flanking region of the *SOD1* gene; respectively. These variants are in strong linkage disequilibrium (r^2^ = 1.0, D′=1.0). The *SOD1* rs2070424 variant has been associated with reduced *SOD1* mRNA expression in immortalized lymphoblastoid cells lines and with decreased SOD1 enzymatic activity (Kase et al., 2012). Although the precise mechanism underlying the altered SOD1 activity remains unclear, regulatory annotation from the FORGEdb database assigns rs2070424a high regulatory potential score (9/10) and indicates overlap with the transcription factor binding motif Hoxa7_3750.1 which is implicated in the regulation of gene expression, morphogenesis and cellular differentiation (FORGEdb - Functional SNP, n.d.). Collectively, these observations suggest that the association between *SOD1* rs2070424 and SNIUAA patients, as well as its impact in SOD activity, is more consistent with a modulatory regulatory effect rather than a direct causative role.

The study has several limitations that should be considered when interpreting the findings. First, the functional assays were performed using serum⍰based measurements, which may not accurately reflect tissue⍰specific or cell⍰type–specific enzymatic activity, and cannot capture isoform⍰specific or substrate⍰dependent effects. Second, the sample sizes available for the functional analyses were relatively small, reducing statistical power and limiting the ability to detect subtle genotype–phenotype relationships. Third, the genetic strategy relied on a candidate⍰gene approach, which, although hypothesis⍰driven, may have missed additional loci contributing to NSAID hypersensitivity; accordingly, several associations observed in the discovery phase did not replicate in the validation cohort. Moreover, NSAID hypersensitivity is a multifactorial condition influenced by environmental, immunological, and epigenetic factors that were not assessed in this work. Finally, the study population consisted exclusively of Caucasian Spanish individuals, which may restrict the generalizability of the results to other ethnic groups and reinforces the need for replication in diverse populations.

The main strengths of this study stem from its comprehensive and integrative design. It leverages a large, well⍰characterized cohort and a rigorous two⍰phase strategy—discovery followed by independent replication—to enhance the robustness and reproducibility of the genetic associations identified. The incorporation of functional enzymatic assays provides biological validation of key findings, moving beyond statistical correlation and strengthening mechanistic interpretation. By examining both cross⍰reactive NSAID hypersensitivity (CR⍰NSAIDs) and selective reactions (SNIUAA), the study offers insight into shared and phenotype⍰specific pathways. Its focus on oxidative stress–related mechanisms introduces an innovative dimension to the field, expanding current understanding beyond COX⍰dependent pathways and classical immune activation. Finally, the application of rigorous statistical methodology, including correction for multiple testing and exploratory interaction modeling, supports the reliability and interpretability of the results and allows identification of complex genetic patterns.

In conclusion, this study demonstrates that genetic variation in oxidative⍰stress and immune⍰regulatory pathways significantly modulates susceptibility to NSAID hypersensitivity. *GSTM5* rs11101989 emerged as the most consistent marker for cross⍰reactive intolerance, while variants in *SOD1, AKR1C3*, and *IL4R* were associated with selective reactions, with functional assays confirming reduced GST and SOD activity in risk⍰genotype carriers. These results highlight redox imbalance as an important modulatory mechanism rather than a direct causal driver of disease. Clinically, targeted genotyping of these variants may enhance patient stratification and support safer NSAID prescribing. Future work should validate these biomarkers in larger, ethnically diverse cohorts and evaluate their practical utility in precision⍰guided therapy.

## 4. Innovation

NSAID hypersensitivity lacks reliable biomarkers, and although oxidative-stress mechanisms have been proposed, prior studies have offered limited genetic or functional validation. This work advances the field by integrating large-scale association analyses with enzymatic activity measurements, demonstrating that *GSTM5* rs11101989 and a *GSTZ1* haplotype confer risk for cross-reactive intolerance through reduced GST activity, while *SOD1* rs2070424/rs2833483 elevate susceptibility to SNIUAA via impaired SOD function (Figures 2–4). These genotype– phenotype links provide the first mechanistic, functionally validated redox-based biomarkers to improve risk stratification in NSAID hypersensitivity (Figure 1).

## 5. Materials and methods

### 5.1 Study population

The study population comprised 1,265 individuals, all of whom were Caucasian Spanish participants. The cohort included 543 healthy controls with no history of DHRs and confirmed tolerance to NSAIDs. These controls were recruited from among students and staff at the University of Extremadura (Cáceres, Spain). In addition, 722 patients with confirmed HRs to NSAIDs. This cohort of patients was composed of 305 SNIUAA patients, 56 N-ERD patients and 361 NIUAA patients. Patients were recruited and clinically evaluated at the Allergy Units of University Hospital (Badajoz, Spain) and Infanta Leonor University Hospital (Madrid, Spain), in accordance with previously established diagnostic protocols (Amo et al., 2025; Pérez-Sánchez et al., 2020). All participants underwent comprehensive assessments, including determination of specific IgE, skin prick testing, intradermal testing, and/or oral drug provocation testing. All participants provided signed informed consent prior to inclusion. The study adhered to the principles outlined in the Declaration of Helsinki and subsequent amendments and was approved by the Ethics Committees of both participating institutions. Demographic and clinical characteristics of the study population are summarized in Table 1. The investigation was conducted in two distinct phases.

### 5.2 Discovery phase

For this phase of the study, a subset of 621 participants was selected. This group consisted of 214 healthy controls with no history of DHRs and confirmed tolerance to NSAIDs, as well as 201 SNIUAA patients, 25 N-ERD patients and 181 NIUAA patients. The characteristics of these individuals are summarized in Table 1.

Genomic DNA was isolated from leukocytes and purified according to standard procedures. From a genetic standpoint, we have recently evaluated the role of redox homeostasis in the mechanisms underlying HRs to NSAIDs and β-lactam antibiotics (Ayuso et al., 2021). In that review, available evidence on associations between single-nucleotide variants (SNVs) in genes encoding redox-related enzymes and susceptibility to drug-induced HRs was systematically assessed (Ayuso et al., 2021). The accumulated data suggested that variants in *GSTM1, GSTP1, GSTT1, TXNRD1, SOD1*, and *SOD2* may contribute to the pathophysiology of these reactions, with the strongest support reported for the *GSTM1* null genotype and the *SOD2* rs4880 polymorphism (Ayuso et al., 2021). Building on these observations, we further investigated additional SNVs in key genes involved in redox regulation that may also influence the development of HRs to NSAIDs (Figure S1, Supplementary Material). SNVs were selected based on their minor allele frequency in the target population (MAF > 0.01), prior implication in pharmacogenetic studies focused on HRs to NSAIDs and predicted functional relevance. The genes examined included members of the aldo–keto reductase family (*AKR1B10, AKR1C1, AKR1C2, AKR1C3, AKR1C4, AKR1E2*) (Guo et al., 2024; Shiiba et al., 2017; Skarydova et al., 2013), catalase (*CAT*) (Saify et al., 2016), glutathione S-transferases (*GSTM1, GSTM3, GSTM5, GSTO1, GSTO2, GSTP1, GSTT1, GSTT2, GSTZ1*) (Agúndez et al., 2011; Alshabeeb et al., 2020; Dragovic et al., 2014; Krishnamurthy et al., 2024; Langaee et al., 2015; Li et al., 2025; Polimanti et al., 2010; Simeunovic et al., 2019), the peroxiredoxin system (PRDX6) (Seibold et al., 2013), superoxide dismutase (*SOD1, SOD2, SOD3*) (Kase et al., 2012; Lucena et al., 2010; Wu et al., 2019), thioredoxin reductases (*TXNRD1, TXNRD2*) (Korobeinikova et al., 2021; Kwon et al., 2012), and cytokine-related genes (*IL10, IL18, IL4R, TGFB1, TNF*) (Bottema et al., 2010; Dabhi and Mistry, 2014; Jiménez-Sánchez et al., 2025; Resende et al., 2017). Genotyping was conducted in triplicate using SNP TaqMan ® assays (Life Technologies S.A., Alcobendas, Madrid, Spain), following the manufacturer’s recommendations and previously published methodologies(Amo et al., 2025). Details of the SNVs analyzed are shown in Table S1.

### 5.3 Replication analyses

This phase involved a cohort of 644 individuals who had not participated in the discovery stage. The sample included 329 healthy controls without a history of DHRs and with confirmed tolerance to NSAIDs, along with 104 SNIUAA patients, 31 N-ERD patients and 180 NIUAA patients. Details of the participants are presented in Table 1.

The SNVs were genotyped in triplicate using SNP TaqMan® assays (Life Technologies S.A., Alcobendas, Madrid, Spain). The SNVs analyzed corresponded to those identified through univariable logistic regression from the groups assessed in the discovery phase (Table S2), following the manufacturer’s instructions and as previously described (Amo et al., 2025).

### 5.4 Functional studies

The activities of redox enzymes, including GSTs, AKRs and SODs were also analysed in serum samples. GST total activity was measured using a Glutathione S-transferase assay kit (#703302 Cayman Chemical Company, Ann Arbor, MI, USA) according to the manufacturer’s instructions. Absorbance was measured at 340Lnm with a Varioskan LUX microplate reader (Thermo Fisher Scientific, Waltham, MA, USA). Total AKR activity was quantified using an Aldo-Keto reductase assay kit (#abx294427 Abbexa Ltd, Cambridge, England, UK) following the manufacturer’s protocol with calibration performed using standard solutions supplied in the kit. Absorbance was recorded at 450Lnm using the same microplate reader. Additionally, total SOD activity was determined using a Superoxide Dismutase assay kit (#706002 Cayman Chemical Company, Ann Arbor, MI, USA) in accordance with the manufacturer’s guidelines. Standard curves were constructed for activity calculation, and absorbance readings were obtained at 450Lnm using the same microplate reader.

### 5.5 Statistical analysis

Quantitative variables were expressed as means accompanied by standard deviations, whereas categorical data were presented as frequencies. Differences between groups were assessed using the chi-square test for sex and the Kruskal-Wallis test for age and enzymatic activities conducted with IBM SPSS Statistics for Windows (version 22.0). Allelic and genotypic frequencies, as well as Hardy-Weinberg equilibrium, were assessed using the SNPassoc package in R (GonzáLez et al., 2007). Comparisons between groups were performed using Fisher’s Exact Test (FET) and the Likelihood Ratio Test (LRT). To account for multiple testing, p-values were adjusted using the Benjamini-Hochberg procedure to control the False Discovery Rate (Benjamini et al., 2001). Association between SNVs and clinical traits were quantified using odds ratios (ORs) with 95% confidence intervals (CI), or relative risk (RR) when the variant was absent in the control group. Each SNV was individually examined for its relationship with binary clinical outcomes through logistic regression (IBM SPSS Statistics for Windows, Version 22.0). Statistical significance was defined as a P-value below 0.05. In addition, logistic regression was applied to identify relevant combinations of SNVs associated with case-control status and to assess the importance of these interactions. For each SNV, dominant, codominant, over-dominant, additive and recessive inheritance models were considered. A bagged logic regression approach for classification was implemented, using 100 bootstrap iterations. The importance of SNV interactions identified by the bagged logic regression was evaluated using Pearson’s chi-square statistic. Corresponding p-values were adjusted for multiple comparisons to identify statistically significant interactions and to establish a ranking of interaction importance. All analyses were performed using R software (R Core Development Team et al., 2019). SNV interactions were identified using the logicFS package (Holger Schwender and Tobias Tietz, 2020).

## Data Availability

All data produced in the present study are available upon reasonable request to the authors

## Author contributions

GMJM: Conceptualization, data curation, formal analysis, investigation, methodology, writing-original draft. GTJ and LNS: conceptualization, data curation, investigation. MM: formal analysis. BLN : conceptualization, data curation, investigation, resources, writing-review and editing. AJAG, GME and AP: Conceptualization, supervision, validation, funding acquisition, writing-review and editing.

## Statements and declarations

### Ethical considerations

The protocol for this study was in accordance with the Declaration of Helsinki and its subsequent revisions and was approved by the respective Ethics Committees of the participating Hospitals.

### Consent to participate

Informed consent was obtained from all patients involved in the study. Written informed consent has been obtained from the patients to participate in this study.

### Consent for publication

Informed consent was obtained from all patients involved in the study to publish this paper.

### Data availability statement

The original data contributions presented in the study are publicly available. This data can be found here: Institutional repository DEHESA of University of Extremadura

## Supplemental material

**Figure S1**. Potential role of redox regulation in the development of drug hypersensitivity reactions. Adapted from Antioxidants (Basel). 2021 Feb 15;10(2):294. doi: 10.3390/antiox10020294. Copyright 2021 by the authors. Licensee MDPI, Basel, Switzerland.

**Figure S2**. Effect of *AKR1C3* rs34186955 genotype on the serum AKR activity (mean ± standard error of mean) in SNIUAA patients

**Figure S3**. Effect of *AKR1C3* rs12529 genotype on the serum AKR activity (mean ± standard error of mean) in SNIUAA patients

**Figure S4**. Effect of *AKR1C3* rs34186955 + rs12529 haplotype on the serum AKR activity (mean ± standard error of mean) in SNIUAA patients

