## Supplementary figures and images for "Oxidative stress and genetic susceptibility to cross-reactive and selective NSAID hypersensitivity"

### Figure S1

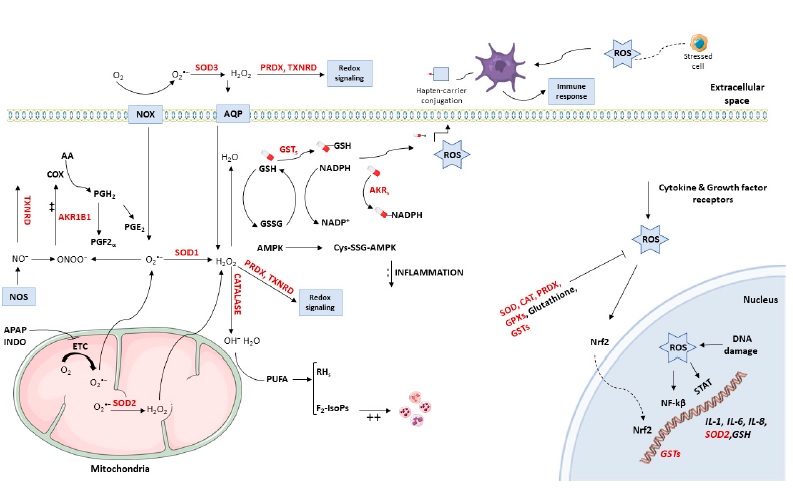
