## Supplementary material for "Oxidative stress and genetic susceptibility to cross-reactive and selective NSAID hypersensitivity": Figure S3

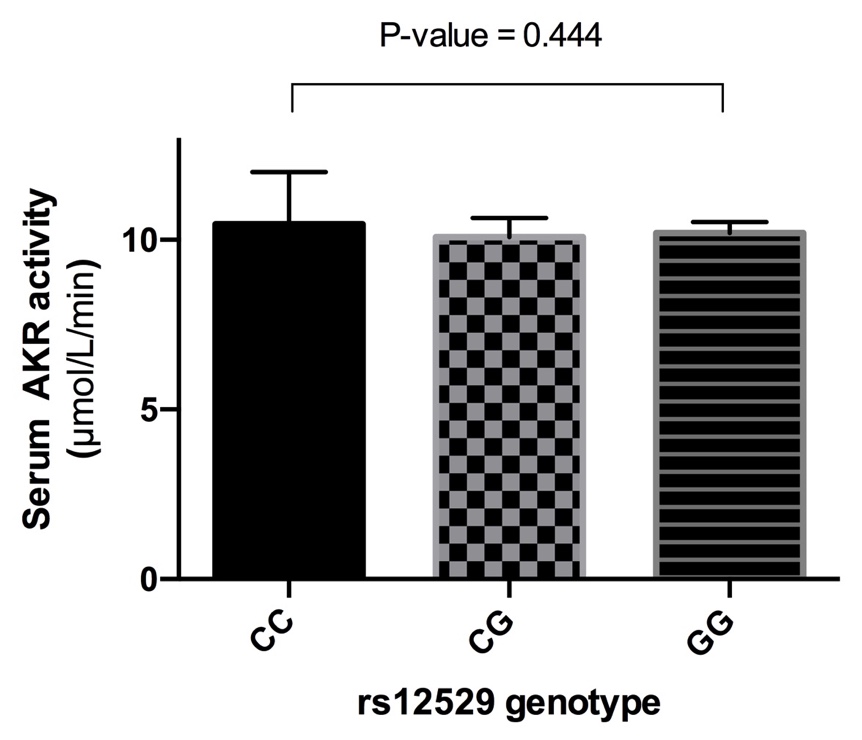

Figure S3. Effect of *AKR1C3* rs12529 genotype on the serum AKR activity (mean ± standard error of mean) in SNIUAA patients
