## Supplementary material for "Oxidative stress and genetic susceptibility to cross-reactive and selective NSAID hypersensitivity": Figure S4

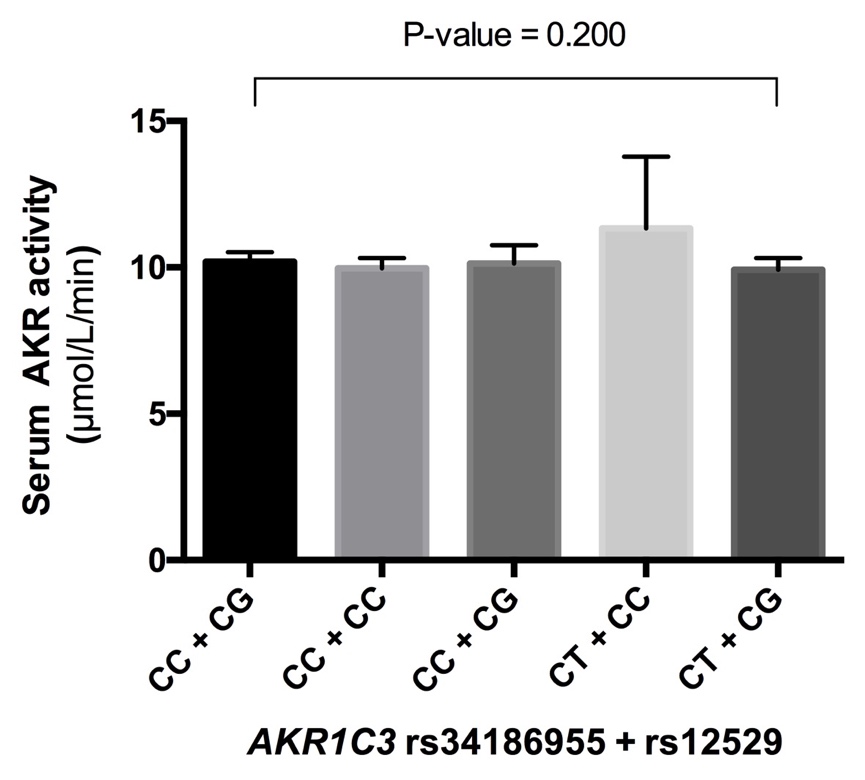

Figure S4. Effect of *AKR1C3* rs34186955 + rs12529 haplotype on the serum AKR activity (mean ± standard error of mean) in SNIUAA patients
